# Prevalence and Integration of symptoms and comorbidities with age of the SARS-CoV-2 patients: a systematic evaluation and meta-analysis

**DOI:** 10.1101/2020.08.19.20177980

**Authors:** Mohammad Meshbahur Rahman, Badhan Bhattacharjee, Zaki Farhana, Mohammad Hamiduzzaman, Muhammad Abdul Baker Chowdhury, Mohammad Sorowar Hossain, Mahbubul H Siddiqee, Md. Ziaul Islam, Enayetur Raheem, Md. Jamal Uddin

**Affiliations:** Biomedical Research Foundation, Dhaka-1230, Bangladesh.; Department of Biotechnology, BRAC University, Dhaka 1212, Bangladesh.; Department of Community Medicine, National Institute of Preventive and Social Medicine, Mohakhali, Dhaka-1212, Bangladesh.; College of Medicine & Public Health, Flinders University, South Australia, Australia.; Department of Emergency Medicine, University of Florida College of Medicine, FL 32608, USA.; Department of Statistics (Biostatistics and Epidemiology), Shahjalal University of Science & Technology, Sylhet-3114, Bangladesh.

**Keywords:** COVID-19 pandemic, Symptoms and Comorbidities, Meta-analysis and Correlation Analysis

## Abstract

**Background:** The COVID-19 affected millions of people, and the patients present a constellation of symptoms and comorbidities. We aimed to estimate the prevalence of reported symptoms and comorbidities and assess the correlation between a series of symptoms and comorbidities and age of the patients’ positive in COVID-19.

**Methods:** We performed a systematic review and meta-analysis [PROSPERO registration: CRD42020182677]. Databases [PubMed, SCOPUS, EMBASE, WHO, Semantic Scholar, and COVID-19 Primer] were searched for clinical studies published from January to April 2020. The pooled prevalence of symptoms and comorbidities were identified using the random effect model, and the multivariable factor analysis was performed to show the correlation between a group of symptoms and comorbidities and age of the COVID-19 patients.

**Findings:** Twenty-nine articles [China (24); Outside of China (5)], with 4,884 COVID-19 patients were included in this study. The meta-analysis investigated 33 symptoms, where fever [84%], cough/dry cough [61%], and fatigue/weakness [42%] were found more prevalent. Out of 43 comorbidities investigated, acute respiratory distress syndrome (ARDS) [61%], hypertension [23%] and diabetes [12%] were the most prevalent comorbid condition. The multivariable factor analysis showed positive association between a group of symptoms and comorbidities, and with the patients’ age. For example, the symptoms comprising fever, dyspnea/shortness of breath, nausea, vomiting, abdominal pain, dizziness, anorexia and pharyngalgia; and the comorbidities including diabetes, hypertension, coronary heart disease, COPD/lung disease and ARDS were positively correlated with the COVID-19 patient’s age.

**Interpretation:** Among 19 symptoms and 11 comorbidities investigated, a group of symptoms and comorbidities were found correlated with the patients infected in COVID-19.

**Summary box**

What is already known?

- Study on the correlation of symptoms and comorbidities with age of the COVID-19 patients are yet to be explored sufficiently.

What are the new findings?

- We investigated all the reported symptoms [33] and comorbidity [43] where fever [84%], cough/dry cough [61%], fatigue/weakness [42%] and dyspnea/shortness of breath [39%] were the most prevalent symptoms, and ARDS [61%], followed by hypertension [23%] and diabetes [12%] were the most prevalent comorbid condition.
- Key findings, a group of symptoms and comorbidities were positively correlated with the COVID-19 patients’ age.
- The symptoms comprising fever, dyspnea/shortness of breath, nausea, vomiting, abdominal pain, dizziness, anorexia and pharyngalgia; and the comorbidities including diabetes, hypertension, coronary heart disease, COPD/lung disease and ARDS were positively associated with the COVID-19 patient’s age.

What do the new findings imply?

- This efforts in determining the correlation of the frequent symptoms and comorbidities will help the clinicians and disease practitioners to implement patient-centred interventions.

## Introduction

The COVID-19 pandemic caused by Severe Acute Respiratory Virus 2 (SARS-CoV-2) is a serious public health crisis in the history of humanity. Originated in Wuhan, China, SARS-CoV-2 has spread to every corner of the world within a few months. As of September 12, 2020, over 28.33 million confirm cases and 911 877 deaths have been reported from over 216 countries.^1^

As the virus is moving fast, various clinical spectrum and differential clinical outcomes are unfolding across different geographic locations. Several symptoms have been reported which includes fever, cough, myalgia, sputum production, headache, hemoptysis, diarrhea, and dyspnea.^2^ The severity of COVID-19 has been reported to be linked with various host factors including diabetes, hypertension, cardiovascular disease, chronic obstructive pulmonary disease (COPD), malignancy, and chronic liver disease.^2^ While susceptibility to COVID-19 covers all age groups, people with compromised immune systems and or having comorbidity are at a higher risk.^3,4^ A few review studies investigated symptoms and comorbidities of the COVID-19 infected patients with a shorter time-frame.^3,6–9^ The mortality rate is high in older COVID-19 patients with organ dysfunctions comprising shock, acute respiratory distress syndrome (ARDS), acute cardiac injury, and acute kidney injury.^5^

However, there is scarce information regarding the relationship among the symptoms, comorbidities and age of the COVID-19 patients. To address this issue, we aimed to estimate all the reported symptoms and comorbidities and assess the correlation between a series of symptoms and comorbidities and age of the COVID-19 infected patients.

## Methods

The PRISMA-P-2009 guidelines was followed in our systematic review and meta-analysis [PROSPERO registration: CRD42020182677].^10^

The major databases, such as PubMed, SCOPUS, EMBASE, WHO, Semantic Scholar, and COVID-19 Primer were searched to include peer-reviewed and pre-proof research articles. The mortality starts falling at the end of April 2020 and we limited our review within initial period to high mortality period to understand the correlation among symptoms, comorbidity, and age of the COVID-19 infected patients. Also, our literature search strategy covered almost hundred percent of the COVID-19 symptoms and comorbidities. Therefore, we restricted our search language in only English literature within the time period January to April 20, 2020.

The search terms used included: “COVID-19” OR “COVID-2019” OR “severe acute respiratory syndrome coronavirus 2” OR “2019-nCoV” OR “2019nCoV” OR “nCoV” OR “SARS-CoV-2” OR “coronavirus” AND “clinical for epidemiological characterization” OR “Symptom” OR “Symptoms” AND “comorbidity” OR “comorbidities”. Some articles were manually retrieved from Google Scholar and other databases. We also searched the reference lists of the selected publications. MMR, BB, and MJU independently screened the titles and abstracts of the articles and checked full-text eligibility [supplementary Table S1].

Research articles were selected if they reported clinical characteristics [both symptoms and comorbidities] of the COVID-19 patients. The inclusion criteria for studies were: clinical investigations or consecutive cases; focused on infected patients; reported at least ten cases and considered all age-groups from any countries. Studies were excluded if they were: grey literature, case report and secondary studies; specific to children or pregnant women; and only reported symptoms or comorbidities. A standardized form was used to extract data from eligible studies. Disagreements were resolved through discussion with co-reviewers. For each study, publication details, research design and the participants’ characteristics with major findings were recorded.

The quality of each study was assessed by ZF using the Joanna Briggs Institute (JBI) guidelines.^11^ A set of eight questions was used for the quality assessment. Random effect model was used to estimate the prevalence of all reported symptoms and comorbidities in the COVID-19 patients. Heterogeneity was assessed using the Cochran Q and the I^2^ statistic.^12,13^ To assess asymmetry and publication bias, we used funnel plot and Egger test [p<0.001] to test the presence of small-study effects. Multivariate analysis [multivariable factor analysis (MFA)] was performed to examine the correlation among symptoms and comorbidities with the patients’ age.^14,15^ All statistical analyses were conducted by Stata version 15 (Stata Corp, College Station, TX) using the metaprop, metabias, metafunnel commands, and R-programming language using the FactoMineR package.

## Results

A total of 799 articles [database: 791, other sources: 8] were retrieved. Of them, 403 articles were removed due to duplication and irrelevance. Furthermore, 303 review articles, editorials, case reports, and irrelevant study populations were excluded. Fifty-three articles were excluded as they failed to meet all inclusion criteria. Finally, eleven articles were excluded due to not peer-reviewed and small sample sizes, resulting in the selection of 29 articles for our review. The PRISMA flow diagram visualizes the screening process of selected studies [Figure 1].

**Figure 1.**
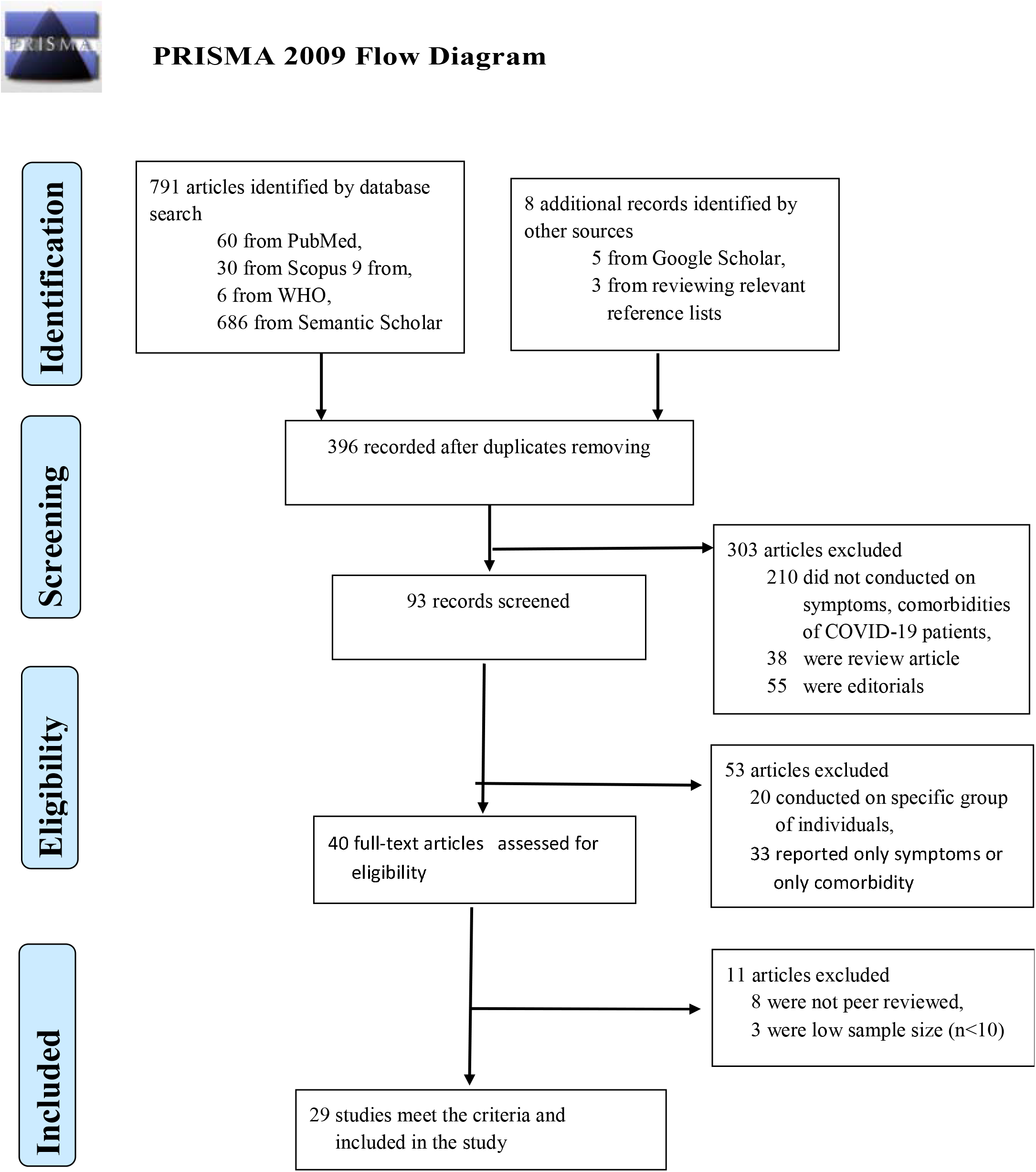
Preferred reporting items for systematic reviews and meta-analyses (PRISMA) flow diagram for the study selection process.

Supplementary table S2 summarizes the characteristics of the selected studies and 83% of selected studies for this meta-analysis were reported from China. Five studies were conducted in the USA, India, Spain, and South Korea. The overall sample size was 4,884 COVID-19 patients, with an age range of 10 to 92 years. Among the patients, 2,675 [55%] were male, and 2,208[45%] were female. The sample size ranged from 12 to 1,099 patients, where most studies (79%) had a retrospective research design.

Altogether, 33 symptoms and 43 comorbidities were found. Almost all the studies reported fever [proportion of patients ranging from 25 to 100%], cough/dry cough [22-92%] and myalgia or muscle ache [3-63%] as common symptoms of COVID-19. Other reported symptoms were: headache [3-66%]; diarrhea [3-48%]; fatigue/weakness [9-85%]; dyspnea/shortness of breath [1-88%]; sputum production or expectoration [4-42%]; vomiting [1-19%]; nausea [4-27%]; chest tightness [7-55%]; and sore throat [5-32%]. Additionally, the less reported symptoms and comorbidities were represented in supplementary Table S2. For the comorbidities, about 93% and 86% of studies reported two comorbidities: diabetes [2 to 35%] and hypertension [8-50%]. Other prevalent comorbidities were chronic obstructive pulmonary disease (COPD)/lung infection [0.2-38%]; cardiovascular disease [5-23%]; chronic liver disease [1-29%]; malignancy [1-7%]; coronary heart disease [1-33%]; cerebrovascular disease [1-19%]; chronic renal disease [1-8%]; chronic kidney disease [1-29%]; and Acute respiratory distress syndrome (ARDS) [17-100%]. The less reported comorbidities were presented in supplementary Table S2.

We meta-analysed 19 symptoms and 11 comorbidities, using a random effect model that were reported in at least five selected articles [Table 1 and supplementary figure S3-S32]. Meta-analysis showed a higher prevalence of fever [pooled prevalence: 84%, 95% confidence interval (CI): 80-88%] and cough/dry cough [61%, 95% CI:55-67%]; followed by fatigue/weakness [42%, 95% CI:34-51%]; dyspnea/shortness of breath [39%, 95% CI:27-51%]; headache and diarrhea [12%, 95% CI:8-17%]; sore throat [15%, 95% CI:11-20%]; myalgia/muscle ache and sputum production/expectoration [24%, 95% CI:18-30%]; rhinorrhea [13%, 95% CI:4-26%]; chest tightness [25%, 95% CI:15-31%]; and anorexia [26%, 95% CI:16-38%]. The less prevalent symptoms were: chest pain [3%], nausea [8%], vomiting [6%], abdominal pain [4%], dizziness [5%], pharyngalgia [7%], and hemoptysis [2%].

**Table 1.** Overall Prevalence summary for clinical symptoms and comorbidities of the COVID-19 patients.

| Clinical Characteristics<br>(Symptoms) | No. Reports | No. Patients | Pooled Prevalence | Test for Heterogeneity |  | Egger's Test |
| --- | --- | --- | --- | --- | --- | --- |
|  |  |  |  | I<sup>2</sup> (%) | P-value |  |
| Fever | 29 (100%) | 4115 | 0.84 (0.80-0.88) | 90.670 | <0.001 | <0.001 |
| Cough / Dry cough | 29(100%) | 3039 | 0.61 (0.55-0.67) | 93.400 | <0.001 | 0.382 |
| Fatigue/Weakness | 21 (72.41%) | 1627 | 0.42 (0.34-0.51) | 96.320 | <0.001 | 0.107 |
| Dyspnoea/Shortness of breath | 18 (62.06%) | 920 | 0.39 (0.27-0.51) | 97.370 | <0.001 | <0.001 |
| Headache | 22 (72.86%) | 448 | 0.12 (0.09-0.16) | 89.980 | <0.001 | 0.109 |
| Diarrhoea | 22 (72.86%) | 474 | 0.12 (0.08-0.17) | 93.720 | <0.001 | 0.004 |
| Sore Throat | 9 (31.03%) | 348 | 0.15 (0.11-0.20) | 84.990 | <0.001 | 0.266 |
| Myalgia/Muscle Ache | 25 (86.20%) | 925 | 0.24 (0.18-0.30) | 95.000 | <0.001 | <0.001 |
| Rhinorrhoea | 5 (17.24%) | 48 | 0.13 (0.04-0.26) | 88.010 | <0.001 | 0.088 |
| Sputum Production/Expectoration | 15 (51.72%) | 1066 | 0.24 (0.19-0.30) | 92.310 | <0.001 | 0.956 |
| Chest tightness | 11 (37.93%) | 462 | 0.25 (0.15-0.31) | 88.440 | <0.001 | 0.527 |
| Chest pain | 5 (17.24%) | 15 | 0.03 (0.01-0.04) | 0.000 | <0.95 | 0.878 |
| Nausea | 12 (41.37%) | 238 | 0.08 (0.04-0.12) | 91.780 | <0.001 | 0.023 |
| Vomiting | 14 (48.27%) | 209 | 0.06 (0.03-0.09) | 88.330 | <0.001 | 0.096 |
| Abdominal Pain | 6 (20.68%) | 42 | 0.04 (0.03-0.06) | 22.890 | <0.26 | 0.431 |
| Dizziness | 6 (20.68%) | 71 | 0.05 (0.03-0.08) | 64.21 | <0.002 | 0.132 |
| Anorexia | 7 (24.13%) | 339 | 0.26 (0.16-0.38) | 94.470 | <0.001 | <0.001 |
| Pharyngalgia | 6 (20.68%) | 86 | 0.07 (0.04-0.13) | 88.030 | <0.001 | 0.017 |
| Haemoptysis | 7 (24.13%) | 47 | 0.02 (0.01-0.04) | 63.480 | <0.01 | 0.005 |
| **Comorbidity** |  |  |  |  |  |  |
| Diabetes | 27 (93.10) | 539 | 0.12 (0.09-0.15) | 83.09 | <0.001 | 0.009 |
| Hypertension | 25 (86.20) | 1096 | 0.23 (0.18-0.28) | 93.24 | <0.001 | 0.149 |
| Cardiovascular Disease | 15 (51.72) | 212 | 0.1 (0.07-0.13) | 73.96 | <0.001 | 0.031 |
| Coronary heart disease | 10 (34.48) | 141 | 0.07 (0.03-0.12) | 92.21 | <0.001 | 0.007 |
| Cerebrovascular disease | 10 (34.48) | 100 | 0.06 (0.02-0.08) | 90.77 | <0.001 | 0.004 |
| COPD/Lung disease | 21 (72.41) | 136 | 0.03 (0.02-0.05) | 86.67 | <0.001 | <0.001 |
| Chronic liver disease | 15 (51.72) | 96 | 0.05 (0.03-0.07) | 78.23 | <0.001 | <0.001 |
| Chronic Renal disease | 9 (31.03) | 32 | 0.01 (0.00-0.03) | 54.63 | <0.001 | 0.003 |
| Chronic Kidney disease | 6 (20.68) | 41 | 0.05 (0.02-0.10) | 86.69 | <0.001 | 0.036 |
| Malignancy | 15 (51.72) | 82 | 0.03 (0.02-0.04) | 68.06 | <0.001 | <0.001 |
| ARDS** | 4 (13.79) | 111 | 0.61 (0.15-0.97) | 98.01 | <0.001 | 0.301 |

\*\* ARDS reported in four studies and we include this study into our analysis because it showed higher prevalence rate.

The most prevalent comorbidities were ARDS [61%, 95% CI: 15-97%], hypertension [23%, 95% CI: 18-28%], and diabetes [12%, 95% CI:9-15%], followed by cardiovascular disease [10%, 95% CI:7-13%]; coronary heart disease [7%, 95% CI:3-12%]; cerebrovascular disease [6%, 95% CI:2-08%]; COPD/lung disease [3%, 95% CI: 02-50%]; chronic liver disease [05%, 95% CI: 03-07%]; chronic Renal disease [0.01%, 95% CI: 00%-03%]; chronic Kidney disease [05%, 95% CI: 02-10%]; and malignancy [03%, 95% CI: 02-04%].

There was a high heterogeneity [I^2^ ranged from 85% to 97%, Cochran Q-statistic p < 0.001] in all the prevalence of symptoms, except chest pain [I^2^ = 0%, Cochran Q-statistic p < 0.95]; abdominal pain [I^2^ = 22.89%, Cochran Q-statistic p < 0.26]; dizziness [I^2^ = 64.21%, Cochran Q-statistic p < 0.002]; and haemoptysis [I^2^ = 63.48%, Cochran Q-statistic p < 0.01]. In the case of comorbidities, the heterogeneity was found higher in almost all the comorbidities [I^2^ ranged from 68.06% to 98.01%, Cochran Q-statistic p < 0.001].

Figure 2 presents the association of symptoms/comorbidities with the patients’ age. Nineteen symptoms and 11 comorbidities were categorized into: symptom group and comorbidity group. The correlation circle represented the between/within-group integration with the patients’ age. The longer vectors indicated more influential than others, and the vectors that were close to each other with the same direction indicated a highly positive association [Figure 2]. Vectors that were the opposite direction showed a negative association, and the vectors with an almost 90-degree angle demonstrated no association. The first principal component showed 31.59% variation and the second one showed 20.45% variation in the dataset.

**Figure 2.**
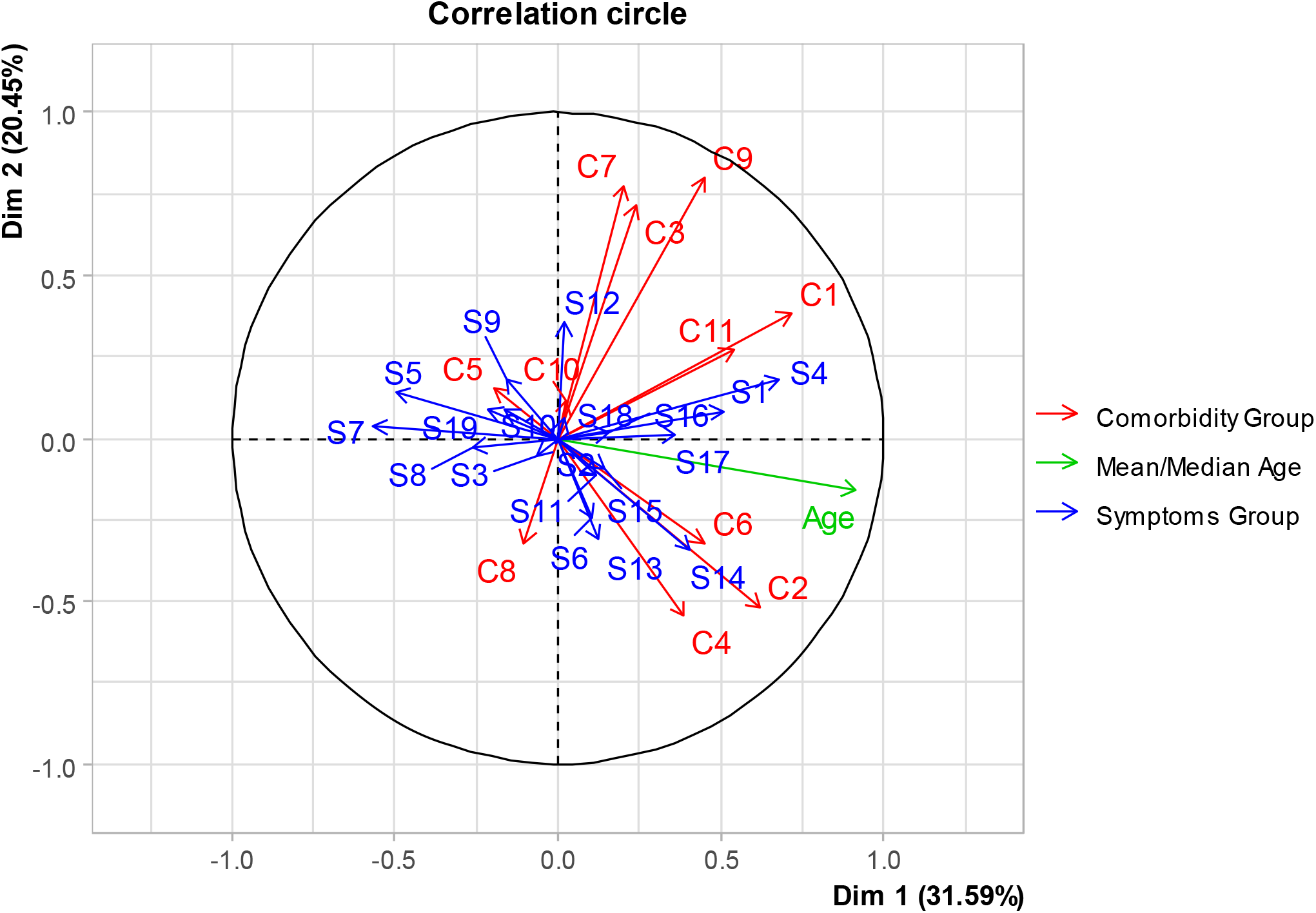
Group Integration [correlation] of symptoms and comorbidities with age of the COVID-19 patients. [**Symptom Group:** S1: Fever, S2: Cough/Dry cough, S3: Fatigue, S4: Dyspnea/Shortness of breath, S5: Headache, S6: Diarrhea, S7: Sore Throat, S8: Myalgia/Muscle Ache, S9: Rhinorrhea, S10: Sputum Production/Expectoration, S11: Chest tightness, S12: Chest pain, S13: Nausea, S14: Vomiting, S15: Abdominal Pain, S16: Dizziness, S17: Anorexia, S18: Pharyngalgia, S19: Haemoptysis. **Comorbidity Group:** C1: Diabetes, C2: Hypertension, C3: Cardiovascular Disease, C4: Coronary heart disease, C5: Cerebrovascular disease, C6: COPD/Lung disease, C7: Chronic liver disease, C8: Chronic Renal disease, C9: Chronic Kidney disease, C10: Malignancy, C11: ARDS.]

In symptom group, fever, dyspnea/shortness of breath, nausea, vomiting, abdominal pain, dizziness, anorexia, and pharyngalgia were found positively associated with the COVID-19 patients’ age. In contrast, sore throat, headache, rhinorrhea, myalgia/muscle ache, fatigue, and hemoptysis were negatively associated with age. Similarly, in the comorbidity group, diabetes, hypertension, coronary heart disease, COPD/lung disease, and ARDS were in the same direction and positively associated with the age of the COVID-19 infected patients. The symptoms like chest tightness/pain and the comorbidities, including chronic liver and kidney diseases, showed no association with the patients’ age.

Considering group integration, the fever, dyspnea/shortness of breath, dizziness, pharyngalgia, and anorexia in the symptom group were positively associated with diabetes, ARDS, and kidney, cardiovascular, and liver diseases in comorbidity group. The symptoms like diarrhea, nausea, vomiting, and abdominal pain were positively associated with hypertension, coronary heart disease, and COPD/lung disease. The symptoms of sore throat, headache, rhinorrhea, myalgia/muscle ache, fatigue, and hemoptysis were positively associated with cerebrovascular disease.

Table 2 summarizes the quality assessment of the selected studies. In 16 [55%] studies, participant recruitment method was appropriate, while the method was unclear in 45% studies. Thirteen [45%] studies had a sample size of more than 100, and about 96% of studies reported the subjects and design in detail. Validated methods were used in all studies, where the measurement was reliable, and the response rate was 100%.

**Table 2.** Quality assessment of the selected studies.

| Authors | Were study participants sampled in an appropriate way? | Was the sample size adequate? | Were the study subjects and the setting described in detail? | Was the data analysis conducted with sufficient coverage of the identified sample? | Were valid methods used for the identification of the condition? | Was the condition measured in a standard, reliable way for all participants? | Was there appropriate statistical analysis? | Was the response rate adequate, and if not, was the low response rate managed appropriately? |
| --- | --- | --- | --- | --- | --- | --- | --- | --- |
| Wan et al.<sup>36</sup> | Yes | Yes | Yes | Yes | Yes | Yes | Yes | Yes |
| Zhang et al.<sup>37</sup> | Yes | Yes | Yes | Yes | Yes | Yes | Yes | Yes |
| Xu et al.<sup>20</sup> | Not Clear | No | Yes | Yes | Yes | Yes | Yes | Yes |
| Zhu et al.<sup>38</sup> | Not Clear | No | Yes | Yes | Yes | Yes | Yes | Yes |
| Chen et al.<sup>39</sup> | Yes | No | Yes | Yes | Yes | Yes | Yes | Yes |
| Liu et al.<sup>40</sup> | Yes | No | Yes | Yes | Yes | Yes | Yes | Yes |
| Chen et al.<sup>22</sup> | Not Clear | Yes | Yes | Yes | Yes | Yes | Yes | Yes |
| Mo et al.<sup>41</sup> | Yes | Yes | Yes | Yes | Yes | Yes | Yes | Yes |
| Liu et al.<sup>21</sup> | Not Clear | Yes | Yes | Yes | Yes | Yes | Yes | Yes |
| Jin et al.<sup>42</sup> | Yes | Yes | Yes | Yes | Yes | Yes | Yes | Yes |
| Wang et al.<sup>16</sup> | Yes | Yes | Yes | Yes | Yes | Yes | Yes | Yes |
| Yuan et al.<sup>23</sup> | Not Clear | No | Yes | Yes | Yes | Yes | Yes | Yes |
| Guan et al.<sup>43</sup> | Yes | Yes | Yes | Yes | Yes | Yes | Yes | Yes |
| Liu et al.<sup>31</sup> | Not Clear | No | Yes | Yes | Yes | Yes | Yes | Yes |
| Zhou et al.<sup>44</sup> | Yes | Yes | Yes | Yes | Yes | Yes | Yes | Yes |
| Huang et al.<sup>2</sup> | Not Clear | No | Yes | Yes | Yes | Yes | Yes | Yes |
| Chen et al.<sup>24</sup> | Yes | No | Yes | Yes | Yes | Yes | Yes | Yes |
| Du et al.<sup>45</sup> | Not clear | No | Yes | Yes | Yes | Yes | Yes | Yes |
| Xu et al.<sup>46</sup> | Yes | No | Yes | Yes | Yes | Yes | Not Clear | Yes |
| Goyal et al.<sup>47</sup> | Not Clear | Yes | Yes | Yes | Yes | Yes | Not Clear | Yes |
| Barrasa et al.<sup>25</sup> | Yes | No | Yes | Yes | Yes | Yes | Yes | Yes |
| Yan et al.<sup>48</sup> | Yes | No | Not Clear | Yes | Yes | Yes | Yes | Yes |
| Gupta et al.<sup>49</sup> | Not Clear | No | Yes | Yes | Yes | Yes | No | Yes |
| Yang et al.<sup>50</sup> | Yes | Yes | Yes | Yes | Yes | Yes | Yes | Yes |
| Han et al.<sup>51</sup> | Not Clear | Yes | Yes | Yes | Yes | Yes | Yes | Yes |
| Kim et al.<sup>52</sup> | Yes | No | Yes | Yes | Yes | Yes | Not Clear | Yes |
| Wang et al.<sup>53</sup> | Yes | Yes | Yes | Yes | Yes | Yes | Yes | Yes |
| Shi et al.<sup>54</sup> | Not Clear | No | Yes | Yes | Yes | Yes | Yes | Yes |
| Yang et al.<sup>26</sup> | Not Clear | No | Yes | Yes | Yes | Yes | Yes | Yes |

The funnel plots for symptoms and comorbidities are presented in a figure [supplementary figure S33-S62]. Funnel plot found the existence of asymmetry and publication bias for all symptoms and comorbidities. The Egger test of symptoms - fever, dyspnea/shortness of breath, diarrhea, myalgia/muscle ache, nausea, anorexia, pharyngalgia, and hemoptysis were found significant [p<0.05], which suggested the presence of small-study effects. The comorbidities-diabetes, cardiovascular disease, cerebrovascular disease, COPD/lung disease, chronic liver disease, chronic renal disease, chronic kidney disease, and malignancy were found significant [p<0.05] by the Egger’s test, that recommended the presence of small-study effects.

## Discussion

Our study aimed to estimate the prevalence of reported symptoms and comorbidities, and assess the correlation between a group of symptoms and comorbidities and age of the patients tested positive for COVID-19. In our systematic review and meta-analysis of 29 studies, involving 4,884 patients, twenty-four [83%] studies performed in China, and five were outside of China. The ratio of infection was reported higher in males than in females [100:82.5], and this result is consistent with previous studies.^2,6,16,17^ It is generally assumed that males are more likely to be infected by bacteria and viruses than females, because of the women’s robust innate and adaptive immune responses.^3,18^ Moreover, the patterns of occupation, social communication, and lifestyle expose males to be exposed more to the agent factors of infectious diseases than in females. The mean or median age of the patients ranged from 40 to 66 years, and we found older people are more susceptible to COVID-19, and our study strongly supports this finding.

We found 33 symptoms and 43 comorbidities in the studies, and our meta-analysis included most reported 19 symptoms and 11 comorbidities. Fever, cough/dry cough, fatigue, dyspnea, anorexia, chest tightness, myalgia, sore throat, rhinorrhea, headache, and diarrhea were highly prevalent symptoms where the others symptoms were found rarely. All studies reported fever [84%] and cough/dry cough [61%] as symptoms consistent with relevant studies across the countries.^19–22^ Previous studies reported hypertension as the most common comorbidity,^3,7,8^, but our study suggests three major comorbidities-acute respiratory distress syndrome (61%), hypertension (23%), and diabetes (12%). Acute respiratory distress syndrome was found a higher prevalence rate [61%] as reported in three studies in China and one in outside China.^23–26^ Our systematic review estimated all the reported symptoms and comorbidities of COVID-19 infected patients. The symptoms like anorexia [26%], chest tightness [25%] and rhinorrhea [13%], and one comorbidity, i.e., acute respiratory distress syndrome[61%] were examined with significant prevalence (p<0.05), but they were under-investigated in the published systematic reviews.^6,7,27,28^

Human aging is associated with declines in adaptive and innate immunity, and it loses the body’s ability to protect against infections.^29,30^ Virologists and clinicians agree that the older adults are more vulnerable to COVID-19, and the patient’s age can strongly be correlated with symptoms and comorbidities.^31–35^ Relating this context, our correlation analysis found a group of symptoms and comorbidities were significantly associated with the in COVID-19 patient’s age. There was a correlation between a cluster of symptoms and comorbidities where symptoms and comorbidities were also found internally correlated. For example, the symptoms including fever, dyspnea/shortness of breath, nausea, vomiting, abdominal pain, dizziness, anorexia and pharyngalgia; and the comorbidities comprising diabetes, hypertension, coronary heart disease, COPD/lung disease and ARDS were positively correlated with the age of COVID-19 patients. Additionally, the fever, dyspnea/shortness of breath, dizziness, pharyngalgia, and anorexia in the symptom group were positively correlated with diabetes, ARDS, and kidney, cardiovascular, and liver diseases in comorbidity group. As a unique initiative, the epitomization of the correlation of symptoms and comorbidities with the age of the of COVID-19 patients which may contribute in designing comprehensive health care to COVID-19 patients.

### Limitation of the Study

During literature search, our selected studies confirmed almost hundred present of the COVID-19 symptoms except the loss of taste or smell and we were limited to only in English texts within the time frame January to April 20, 2020. The majority of the studies were found in China, and only five from other countries. More studies outside of China could add value in prevalence estimation. We found no data for <10 years children and thus, more studies are warranted in the child COVID-19 patients. Lastly, a few studies were found low sample size.

## Conclusion

In conclusion, this systematic review and meta-analysis is the pioneering effort of its kind that reports all frequent symptoms and comorbidities, and determines the correlation with the age of the COVID-19 patients. This review may help the clinicians, health care providers, policymakers to take effective strategies for the management of the COVID-19 patients.

## Supporting information

All Supplemental Table and Figures

## Data Availability

The full list of data and the data entries for all included studies is provided in the manuscript. No additional supporting data is available.

## Acknowledgments

We are thankful to the Health Assistant (Md. Saiful Islam) who supported and provided computer and necessary software during the data analysis.

## Author contributions

MMR and MJU contributed to the design and conceptualization of the study. MMR, BB and ZF contributed to the screening of studies for inclusion and data extraction. MMR, BB, MJU and MABC searched the databases. MMR and ZF contributed to the analysis and interpretation of the data. MMR, BB and MH contributed to the drafting and formatting of the manuscript. MMR, MH, MABC, MSH, MHS, MZI, ER and MJU contributed to the data extraction, writing, and editing of the manuscript. All authors contributed to the reviewing for important intellectual context and approved of the manuscript to be submitted.

## Data availability statement

The full list of data and the data entries for all included studies is provided in the manuscript as a supplementary file. No additional supporting data is available.

## Ethics statement

Not Applicable

