## Supplementary material for "Prevalence and Integration of symptoms and comorbidities with age of the SARS-CoV-2 patients: a systematic evaluation and meta-analysis": All Supplemental Table and Figures

**Supplementary Files**

S1. Search strategy used in different databases

| PubMed | SCOPUS/EMBASE | WHO/Semantic Scholar |
| --- | --- | --- |
| ALL ( "COVID-19" OR "COVID-2019" OR "severe acute respiratory syndrome coronavirus 2" OR "severe acute respiratory syndrome coronavirus 2" OR "2019-nCoV" OR "nCoV" OR "SARS-CoV-2" OR "2019nCoV" OR "coronavirus" ) AND ALL ( "clinical for epidemiological characterization" OR "Symptom" OR "Symptoms" ) AND ALL ( "comorbidity" OR "comorbidities" AND full text[sb] AND ( "2019/12/31"[PDat] : "2020/04/20"[PDat] ) AND Humans[Mesh]. | ALL ( "COVID-19"  OR  "COVID-2019"  OR  "severe acute respiratory syndrome coronavirus 2"  OR  "severe acute respiratory syndrome coronavirus 2"  OR  "2019-nCoV"  OR  "nCoV"  OR  "SARS-CoV-2"  OR  "2019nCoV"  OR  "coronavirus" )  AND  ALL ( "clinical for epidemiological characterization"  OR  "Symptom"  OR  "Symptoms" )  AND  ALL ( "comorbidity"  OR  "comorbidities" )  AND  ( LIMIT-TO ( LANGUAGE ,  "English" ) )  AND  ( LIMIT-TO ( PUBYEAR ,  2020 ) )  AND  ( LIMIT-TO ( DOCTYPE ,  "ar" )  OR  LIMIT-TO ( DOCTYPE ,  "re" ) ) | ("COVID-19" OR "COVID-2019" OR "severe acute respiratory syndrome coronavirus 2" OR "severe acute respiratory syndrome coronavirus 2" OR "2019-nCoV" OR "nCoV" OR "SARS-CoV-2" OR "2019nCoV" OR "coronavirus") AND ("clinical for epidemiological characterization" OR "Symptom" OR "Symptoms") AND ("comorbidity" OR "comorbidities") |

**S2.** Characteristics of studies that evaluated the prevalence and correlation of symptoms and comorbidities of the COVID-19 patients.

| **Authors** | **Publication Date** | **Country & Location** | **Study Design** | **Mean/**  **Median**  **Age** | **Patient No. (n)** | **Male (n)** | **Female (n)** | **Gender** | **Fever** | **Cough / Dry cough** | **Fatigue** | **Dyspnoea/**  **Shortness of breath** | **Headache** | **Diarrhoea** |
| --- | --- | --- | --- | --- | --- | --- | --- | --- | --- | --- | --- | --- | --- | --- |
| Wan et al. | 21.3.2020 | Chongqing, China | N/M | 47 | 135 | 72 | 63 | Both | 0.89 | 0.77 | 0.33 | 0.13 | 0.33 | 0.13 |
| Zhang et al. | 18.2.2020 | Wuhan, China | N/M | 57 | 140 | 71 | 69 | Both | 0.79 | 0.64 | 0.64 | 0.31 | N/M | 0.13 |
| Xu et al. | 19.2.2020 | Zhejiang, China | Retrospective | 41 | 62 | 35 | 27 | Both | 0.77 | 0.81 | 0.52 | N/M | 0.34 | 0.08 |
| Zhu et al. | 13.3.2020 | Anhui, China | Retrospective | 46 | 32 | 15 | 17 | Both | 0.84 | 0.66 | 0.16 | N/M | 0.03 | 0.03 |
| Chen et al. | 16.02.2020 | Wuhan, China | Retrospective | 56 | 21 | 17 | 4 | Both | 0.99 | 0.80 | 0.85 | 0.52 | 0.10 | 0.20 |
| Liu et al. | 9.02.2020 | Shenzhen, China | N/M | 60 | 12 | 8 | 4 | Both | 0.83 | 0.92 | N/M | N/M | N/M | 0.17 |
| Chen et al. | 26.03.2020 | Wuhan, China | Retrospective | 62 | 274 | 171 | 103 | Both | 0.91 | 0.68 | 0.50 | 0.44 | 0.11 | 0.28 |
| Mo et al. | 16.3.2020 | Wuhan, China | Retrospective, single center | 54 | 155 | 86 | 69 | Both | 0.81 | 0.63 | 0.73 | 0.32 | 0.10 | 0.05 |
| Liu et al. | 07.02.2020 | Hubei, China | Retrospective | 57 | 137 | 61 | 76 | Both | 0.82 | 0.48 | 0.32 | 0.19 | 0.10 | 0.08 |
| Jin et al. | 24.03.2020 | Zhejiang, China | Retrospective | 46 | 651 | 331 | 320 | Both | 0.84 | 0.67 | 0.18 | N/M | 0.10 | N/M |
| Wang et al. | 17.03.2020 | Wuhan, China | Retrospective, single center | 56 | 138 | 75 | 63 | Both | 0.99 | 0.59 | 0.70 | 0.31 | 0.07 | 0.10 |
| Yuan et al. | 19.03.2020 | Hubei, China | Retrospective | 60 | 27 | 12 | 15 | Both | 0.78 | 0.59 | N/M | 0.41 | N/M | N/M |
| Guan et al. | 28.02.2020 | China (30 provinces) | Cohort | 47 | 1099 | 639 | 460 | Both | 0.89 | 0.68 | 0.38 | N/M | 0.14 | 0.04 |
| Liu et al. | 27.03.2020 | Hainan, China | Retrospective | 68 | 56 | 31 | 25 | Both | 0.76 | 0.36 | 0.09 | N/M | N/M | N/M |
| Zhou et al. | 12.03.2020 | Wuhan, China | N/M | 51 | 254 | 115 | 139 | Both | 0.84 | 0.39 | 0.52 | 0.04 | 0.11 | N/M |
| Huang et al. | 24.01.2019 | Wuhan, China | Cohort | 49 | 41 | 30 | 11 | Both | 0.98 | 0.76 | 0.44 | 0.55 | 0.08 | 0.03 |
| Chen et al. | 29.01.2019 | Wuhan, China | Retrospective, single center | 55.5 | 99 | 67 | 32 | Both | 0.83 | 0.82 | N/M | N/M | 0.08 | 0.02 |
| Du et al. | 3.04.2020 | Wuhan, China | Retrospective | 66 | 85 | 62 | 23 | Both | 0.92 | 0.22 | 0.59 | 0.71 | 0.05 | 0.19 |
| Xu et al. | 28.02.2020 | Guangzhou, China | N/M | 50 | 90 | 39 | 51 | Both | 0.78 | 0.63 | 0.21 | N/M | 0.04 | 0.06 |
| Goyal et al. | 17.04.2020 | New York, USA | Retrospective | 62 | 393 | 238 | 155 | Both | 0.77 | 0.79 | N/M | 0.57 | N/M | 0.24 |
| Barrasa et al. | 1.04.2020 | Vitoria, Spain | N/M | 63 | 48 | 27 | 21 | Both | 1.00 | 0.73 | N/M | 0.88 | N/M | N/M |
| Yan et al. | 12.4.2020 | USA | Cross sectional | 48.5 | 59 | 29 | 29 | Both | 0.70 | 0.66 | 0.81 | 0.54 | 0.66 | 0.48 |
| Gupta et al. | 6.04.2020 | New Delhi, India | Retrospective, Observational | 40 | 21 | 14 | 7 | Both | 0.43 | 0.43 | N/M | N/M | 0.14 | N/M |
| Yang et al. | 21.02.2020 | Wenzhou, China | Retrospective cohort | 45 | 149 | 81 | 68 | Both | 0.77 | 0.58 | N/M | 0.01 | 0.09 | 0.07 |
| Han et al. | 15.04.2020 | Wuhan, China | Retrospective | 62.5 | 206 | 91 | 115 | Both | 0.67 | 0.26 | 0.45 | N/M | N/M | 0.33 |
| Kim et al. | 6.04.2020 | South Korea | Cohort | 40 | 28 | 15 | 13 | Both | 0.25 | 0.29 | 0.11 | N/M | 0.25 | 0.11 |
| Wang et al. | 15.03.2020 | Wuhan, China | Retrospective, single-centre | 69 | 339 | 166 | 173 | Both | 0.92 | 0.53 | 0.40 | 0.41 | 0.04 | 0.13 |
| Shi et al. | 24.02.2020 | Wuhan, China | Retrospective | 49.5 | 81 | 42 | 39 | Both | 0.73 | 0.59 | 0.09 | 0.42 | 0.06 | 0.04 |
| Yang et al. | 21.02.2020 | Wuhan, China | Retrospective, single-centre, Observational | 60 | 52 | 35 | 17 | Both | 0.98 | 0.77 | N/M | 0.64 | 0.06 | N/M |

N/M: Not Mentioned

**S2 (Continued)**. Characteristics of studies that evaluated the prevalence and correlation of symptoms and comorbidities of the COVID-19 patients.

| Authors | Sore Throat | Myalgia/  Muscle Ache | Rhinorrhea | Cough/  Sputum Production | Chest tightness | Chest pain | Nausea | Vomiting | Abdominal Pain | Dizziness | Anorexia | Pharyngalgia | Hemoptysis | Others | No. of Symptoms |
| --- | --- | --- | --- | --- | --- | --- | --- | --- | --- | --- | --- | --- | --- | --- | --- |
| Wan et al. | N/M | 0.33 | N/M | 0.09 | N/M | N/M | N/M | N/M | N/M | N/M | N/M | 0.18 | 0.03 | Loss of appetite-4.4%, Palpitation-3.7%, Retching-3% | 13 |
| Zhang et al. | N/M | N/M | N/M | N/M | 0.31 | N/M | 0.17 | N/M | 0.06 | N/M | 0.17 | N/M | N/M | N/M | 9 |
| Xu et al. | N/M | 0.52 | N/M | N/M | N/M | N/M | N/M | N/M | N/M | N/M | N/M | N/M | 0.03 | N/M | 7 |
| Zhu et al. | N/M | 0.16 | N/M | 0.16 | 0.09 | N/M | N/M | N/M | N/M | N/M | N/M | N/M | N/M | N/M | 8 |
| Chen et al. | N/M | 0.40 | N/M | N/M | 0.55 | N/M | N/M | N/M | N/M | N/M | N/M | N/M | N/M | N/M | 8 |
| Liu et al. | N/M | 0.33 | N/M | N/M | N/M | N/M | 0.17 | 0.17 | N/M | N/M | N/M | N/M |  | Chill- 42% | 7 |
| Chen et al. | N/M | 0.22 | N/M | 0.30 | 0.38 | N/M | 0.09 | 0.06 | 0.07 | 0.08 | 0.24 | 0.04 | 0.03 | N/M | 16 |
| Mo et al. | N/M | 0.61 | N/M | N/M | 0.39 | 0.04 | 0.04 | 0.04 | 0.02 | 0.02 | 0.32 | N/M |  | N/M | 14 |
| Liu et al. | N/M | 0.32 | N/M | 0.04 | N/M | N/M | N/M | N/M | N/M | N/M | N/M | N/M | 0.05 | Heart palpitation-7% | 10 |
| Jin et al. | 0.15 | 0.11 | N/M | 0.35 | N/M | N/M | N/M | N/M | N/M | N/M | N/M | N/M | 0.02 | Nasal Obstruction-6%, | 9 |
| Wang et al. | N/M | 0.35 | N/M | 0.27 | N/M | N/M | 0.10 | 0.04 | N/M | 0.09 | 0.40 | 0.17 | N/M | N/M | 13 |
| Yuan et al. | N/M | 0.11 | N/M | N/M | N/M | N/M | N/M | N/M | N/M | N/M | N/M | N/M | N/M | N/M | 4 |
| Guan et al. | 0.14 | 0.15 | N/M | 0.34 | N/M | N/M | 0.05 | 0.05 | N/M | N/M | N/M | N/M | 0.01 | Conjunctival congestion-1%, Nasal congestion-5%, Chils-11.5%, Throat congestion-2%, Tonsil sweling-2%, Rash-0.2% | 17 |
| Liu et al. | N/M | N/M | N/M | N/M | 0.07 | N/M | N/M | 0.17 | N/M | N/M | N/M | N/M | N/M | Nasal congestion-5%, | 6 |
| Zhou et al. | 0.06 | 0.34 | N/M | 0.42 | 0.26 | N/M | N/M | N/M | N/M | 0.07 | N/M | N/M | N/M | N/M | 10 |
| Huang et al. | N/M | 0.44 | N/M | 0.28 | N/M | N/M | N/M | N/M | N/M | N/M | N/M | N/M | 0.05 | N/M | 9 |
| Chen et al. | 0.05 | 0.11 | 0.04 | N/M | N/M | 0.02 | 0.01 | 0.01 | N/M | N/M | N/M | N/M | N/M | N/M | 10 |
| Du et al. | N/M | 0.17 | N/M | 0.38 | N/M | 0.02 | N/M | 0.05 | 0.04 | N/M | 0.57 | 0.02 | N/M | N/M | 14 |
| Xu et al. | 0.26 | 0.28 | N/M | 0.12 | N/M | N/M | 0.06 | 0.02 | N/M | N/M | N/M | N/M | N/M | Chills-7% | 11 |
| Goyal et al. | N/M | 0.19 | N/M | N/M | N/M | N/M | 0.19 | 0.19 | N/M | N/M | N/M | N/M | N/M | N/M | 7 |
| Barrasa et al. | N/M | 0.04 | N/M | N/M | N/M | N/M | N/M | N/M | N/M | N/M | N/M | N/M | N/M | Malaise-44% | 5 |
| Yan et al. | 0.32 | 0.63 | 0.31 | N/M | N/M | N/M | 0.27 | N/M | N/M | N/M | N/M | N/M | N/M | Nasal obstruction 47.5%, Anosmia 68%, Ageusia 71% | 13 |
| Gupta et al. | 0.24 | N/M | N/M | N/M | N/M | N/M | N/M | N/M | N/M | N/M | N/M | N/M | N/M | N/M | 5 |
| Yang et al. | 0.14 | 0.03 | N/M | 0.32 | 0.10 | 0.03 | 0.01 | 0.01 | N/M | N/M | N/M | N/M | N/M | Chill-14%, Snotty-3% | 14 |
| Han et al. | N/M | 0.21 | N/M | N/M | 0.24 | N/M | N/M | 0.12 | 0.04 | N/M | N/M | 0.06 | N/M | Poor apitite-34% | 11 |
| Kim et al. | 0.29 | 0.25 | 0.07 | 0.21 | N/M | N/M | N/M | N/M | 0.04 | N/M | N/M | N/M | N/M | N/M | 10 |
| Wang et al. | N/M | 0.05 | N/M | 0.28 | 0.26 | N/M | 0.04 | N/M | N/M | 0.04 | 0.28 | 0.04 | N/M | N/M | 13 |
| Shi et al. | N/M | N/M | 0.26 | 0.19 | 0.22 | N/M | N/M | 0.05 | N/M | 0.02 | 0.01 | N/M | N/M | N/M | 12 |
| Yang et al. | N/M | 0.12 | 0.06 | N/M | N/M | 0.02 | N/M | 0.04 | N/M | N/M | N/M | N/M | N/M | Malaise-35%, Arthralgia-2% | 10 |

N/M: Not Mentioned

**S2 (Continued)**. Characteristics of studies that evaluated the prevalence and correlation of symptoms and comorbidities in the nCOVID-19 patients.

| **Authors** | **Diabetes** | **Hypertension** | **Cardiovascular Disease** | **Coronary heart disease** | **Cerebrovascular disease** | **COPD/Lung disease** | **Chronic liver disease** | **Chronic Renal disease** | **Chronic Kidney disease** | **Malignancy** | **ARDS** | **Others** | **No. of Comorbidities** |
| --- | --- | --- | --- | --- | --- | --- | --- | --- | --- | --- | --- | --- | --- |
| Wan et al. | 0.09 | 0.10 | 0.05 | N/M | N/M | 0.007 | 0.02 | N/M | N/M | 0.03 | N/M | N/M | 6 |
| Zhang et al. | 0.12 | 0.30 | N/M | 0.05 | N/M | 0.014 | 0.06 | 0.01 | N/M | N/M | N/M | Gastric ulcer & Hyperlepedemia-5%, Thyroid disease-3.6%, Urolithiasis-2.1%, Arrhythmia-3.6%, Cholilithiasis-4.3% | 12 |
| Xu et al. | 0.02 | 0.08 | N/M | N/M | 0.02 | 0.020 | 0.11 | 0.02 | N/M | N/M | N/M | N/M | 6 |
| Zhu et al. | 0.13 | 0.22 | N/M | 0.06 | 0.03 | 0.060 | 0.06 | 0.03 | N/M | N/M | N/M | Mental disorder- 3%, Tumor-6% | 9 |
| Chen et al. | 0.14 | 0.24 | N/M | N/M | N/M | N/M | N/M | N/M | N/M | N/M | N/M | N/M | 2 |
| Liu et al. | 0.08 | 0.25 | N/M | 0.33 | N/M | 0.080 | N/M | 0.08 | N/M | N/M | N/M | Bacterial co infection- 17%, Pneumonia-100%, | 7 |
| Chen et al. | 0.17 | 0.34 | 0.08 | N/M | 0.01 | 0.070 | N/M | N/M | 0.01 | 0.03 | N/M | HBV infection-4%, Metabolic arthritis & Autoimmune disease & Gastro Intestinal disease- 1% | 11 |
| Mo et al. | 0.10 | 0.24 | 0.10 | N/M | 0.05 | 0.030 | 0.05 | 0.04 | N/M | 0.05 | N/M | Tuberculosis-2%, HIV-1% | 10 |
| Liu et al. | 0.10 | 0.10 | 0.07 | N/M | N/M | 0.015 | N/M | N/M | N/M | 0.02 | N/M | N/M | 5 |
| Jin et al. | 0.07 | 0.15 | N/M | 0.01 | N/M | 0.002 | 0.04 | 0.01 | N/M | 0.01 | N/M | Immunosuppression-0.17%, | 8 |
| Wang et al. | 0.10 | 0.31 | 0.15 | N/M | 0.05 | 0.030 | 0.03 | N/M | 0.03 | 0.07 | N/M | HIV infection-1% | 9 |
| Yuan et al. | 0.22 | 0.19 | 0.11 | N/M | N/M | N/M | N/M | N/M | N/M | N/M | 0.41 | Tumor & Cerebral infraction & Chronic gastric- 4% | 9 |
| Guan et al. | 0.07 | 0.15 | N/M | 0.03 | 0.01 | 0.010 | N/M | 0.01 | N/M | 0.01 | N/M | Hepatitis B Infection-2%, Immune deficiency-0.2%, | 9 |
| Liu et al. | 0.07 | 0.17 | N/M | 0.11 | N/M | N/M | 0.06 | N/M | 0.03 | N/M | N/M | Persistent arterial fibrilliation-6%, | 6 |
| Zhou et al. | 0.10 | 0.25 | 0.05 | 0.07 | N/M | 0.020 | 0.01 |  | N/M | 0.01 | N/M | HIV infection-0.4%, | 8 |
| Huang et al. | 0.20 | 0.15 | 0.15 | N/M | N/M | 0.020 | 0.02 | N/M | N/M | 0.02 | N/M | N/M | 6 |
| Chen et al. | N/M | N/M | N/M | N/M | N/M | N/M | N/M | 0.03 | N/M | N/M | 0.17 | Acute Respiratory Injury-8%, Septic shock-4%, Pneumonia-1% | 5 |
| Du et al. | 0.22 | 0.38 | 0.08 | 0.12 | N/M | 0.020 | 0.06 | N/M | 0.04 | 0.07 | N/M | N/M | 8 |
| Xu et al. | 0.06 | 0.19 | 0.03 | N/M | N/M | 0.010 | N/M | N/M | N/M | 0.02 | N/M | Tuberculosis-2%, | 6 |
| Goyal et al. | 0.25 | 0.50 | N/M | 0.14 | N/M | 0.050 | N/M | N/M | N/M | N/M | N/M | Asthama-12.5%, Obesity-35% | 6 |
| Barrasa et al. | 0.19 | 0.44 | N/M | 0.10 | N/M | 0.380 | N/M | N/M | N/M | N/M | 1.00 | Obesity-48%, Immunosuppression-3% | 7 |
| Yan et al. | 0.09 | 0.14 | 0.05 | N/M | N/M | 0.050 | N/M | N/M | N/M | 0.04 | N/M | Allergic rhinitis-34%, Sinus disease 3% | 7 |
| Gupta et al. | 0.14 | 0.24 | N/M | N/M | N/M | N/M | N/M | N/M | N/M | N/M | N/M | Anxity-5%, Hypothyroidism-5%, Migrane-5%, Obstructive sleep aponea-5% | 6 |
| Yang et al. | N/M | N/M | 0.19 | N/M | 0.19 | N/M | N/M | N/M | N/M |  | N/M | Respiratory system disease - 0.67%, Digestive system disease- 5%, Endocrine disease- 6% | 5 |
| Han et al. | 0.10 | 0.27 | N/M | N/M | 0.08 | 0.040 | N/M | N/M | N/M | N/M | N/M | N/M | 4 |
| Kim et al. | 0.07 | N/M | N/M | N/M |  | N/M | 0.04 |  | N/M | 0.04 | N/M | Obesity-18%, Asthama-4% | 5 |
| Wang et al. | 0.16 | 0.41 | 0.16 | N/M | 0.06 | 0.060 | 0.01 | N/M | 0.04 | 0.04 | N/M | Autoimmune disease-1.5% | 9 |
| Shi et al. | 0.12 | 0.15 | 0.10 | N/M | 0.07 | 0.110 | 0.09 | 0.04 | N/M | 0.05 | N/M | N/M | 8 |
| Yang et al. | 0.35 | N/M | 0.23 | N/M | N/M | N/M | 0.29 | N/M | 0.29 | N/M | 0.67 | Urinary tract infection-2%, Gastrointestinal haemorrhage-4% | 7 |

N/M: Not Mentioned

**Overall Meta-analysis of the clinical symptoms and comorbidities**

**Symptoms**

**S3. Fever**

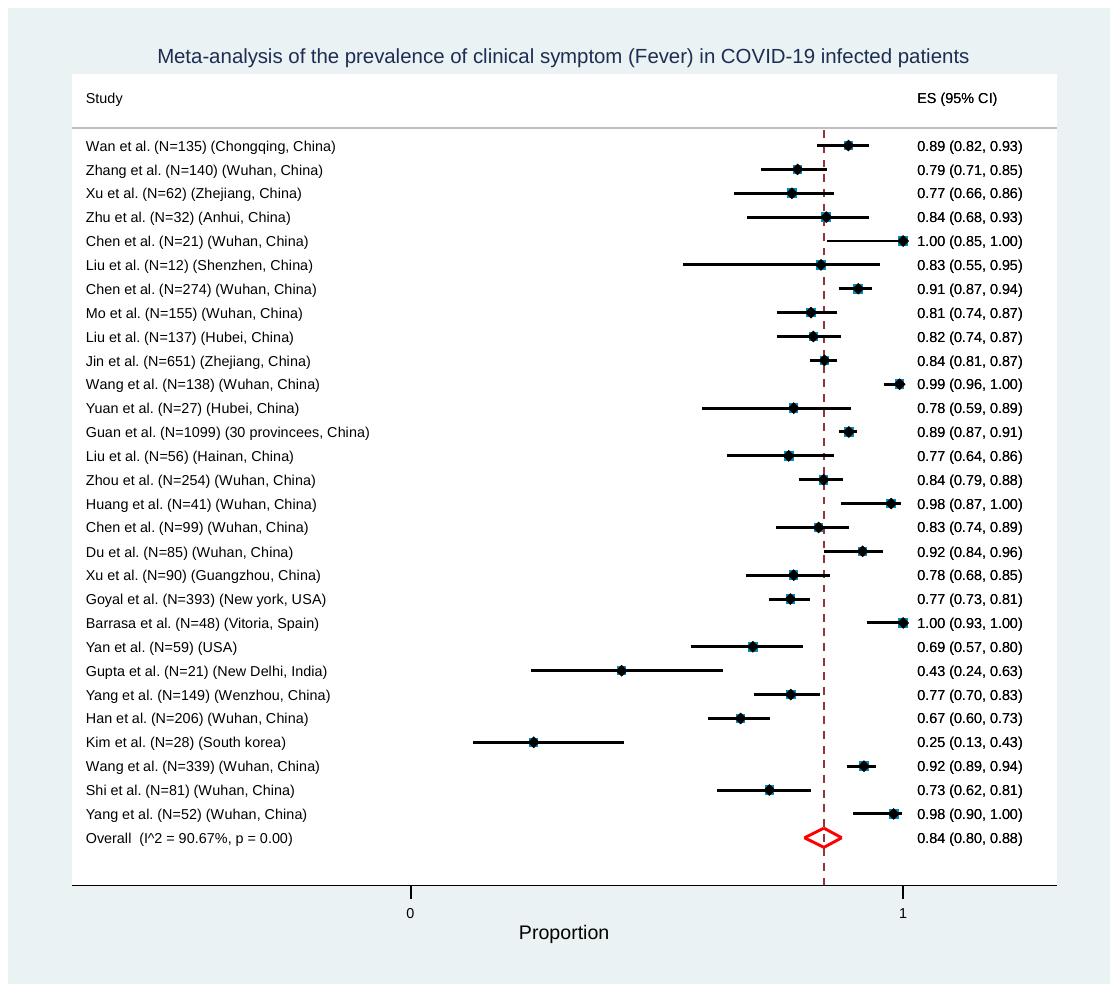

**S4. Cough / Dry Cough**

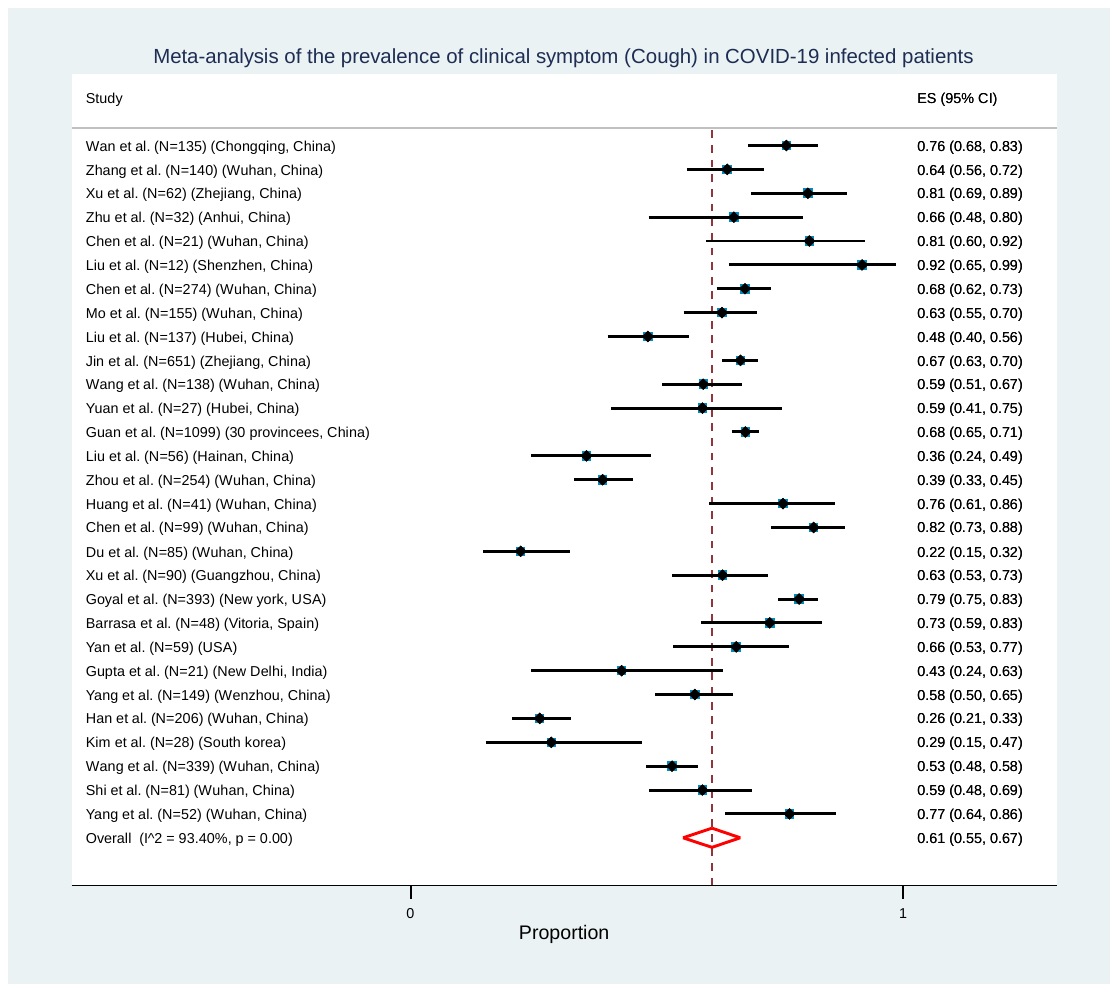

**S5. Fatigue/Weakness**

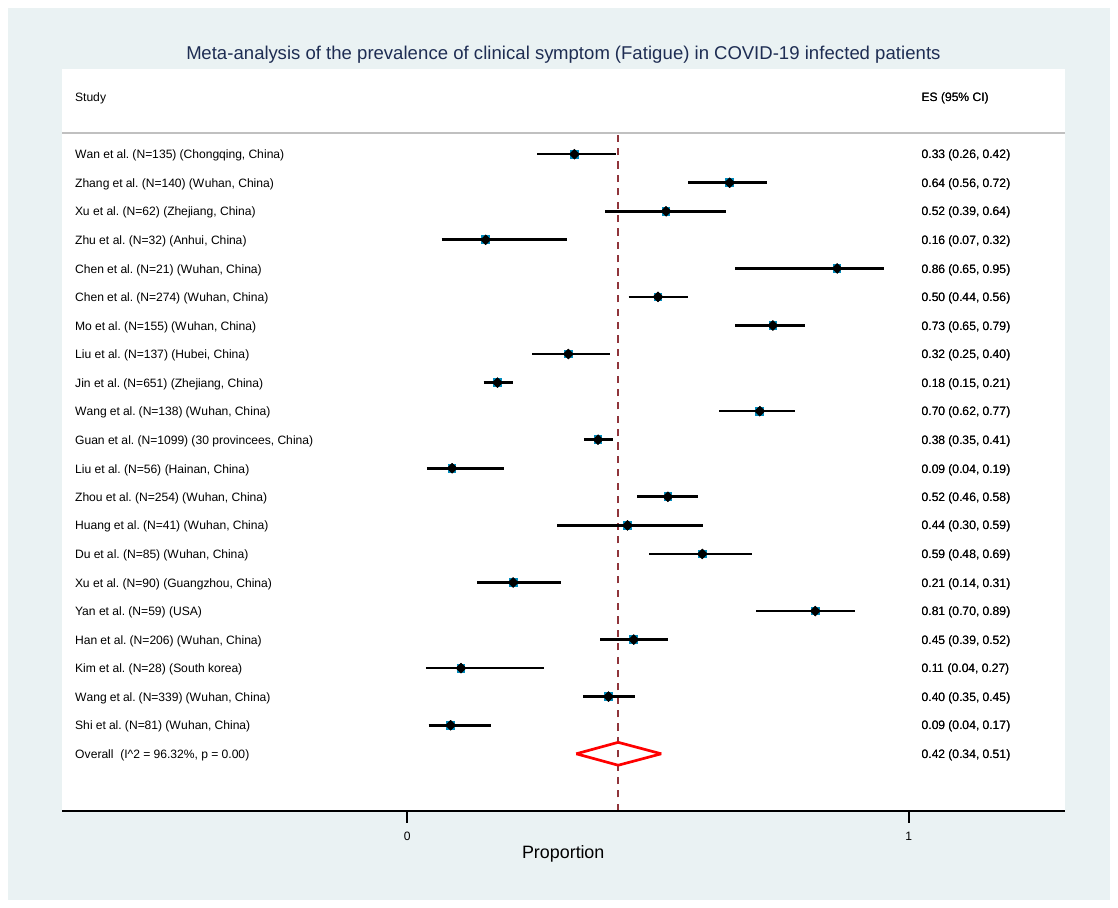

**S6. Dyspnea/Shortness of breath**

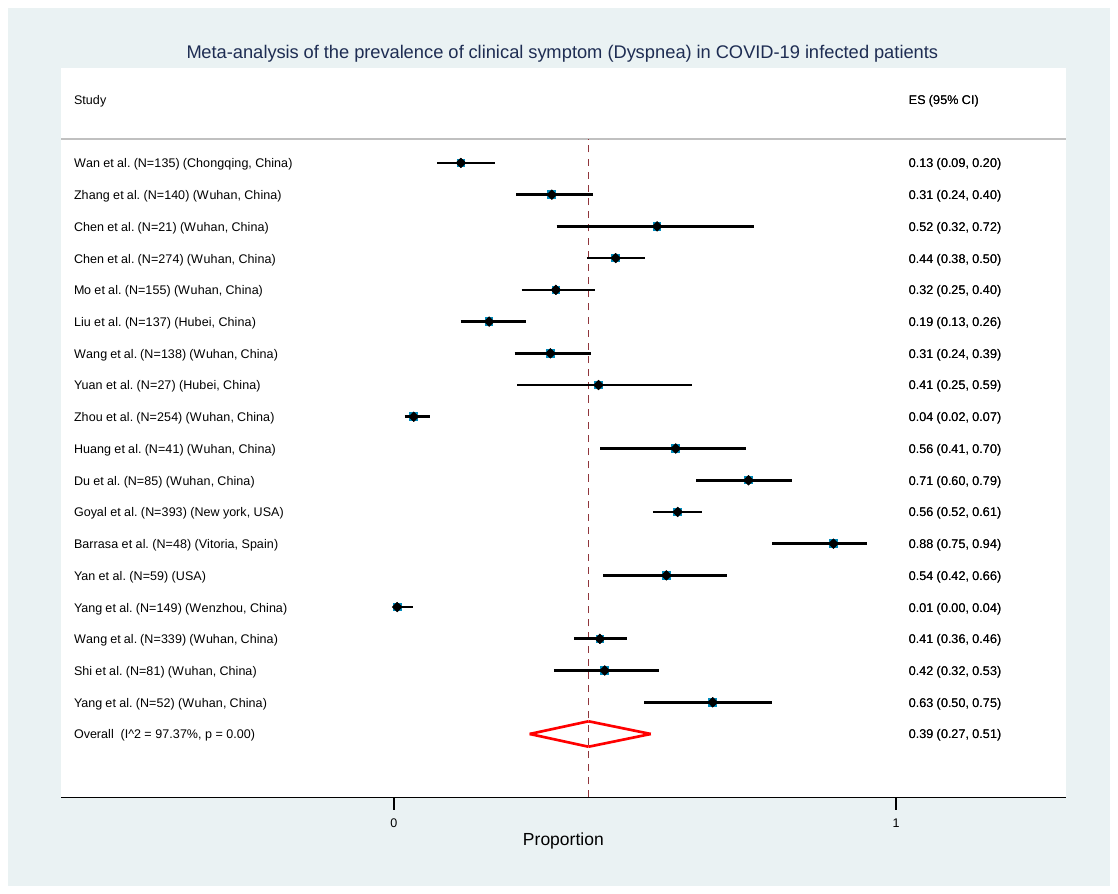

**S7. Headache**

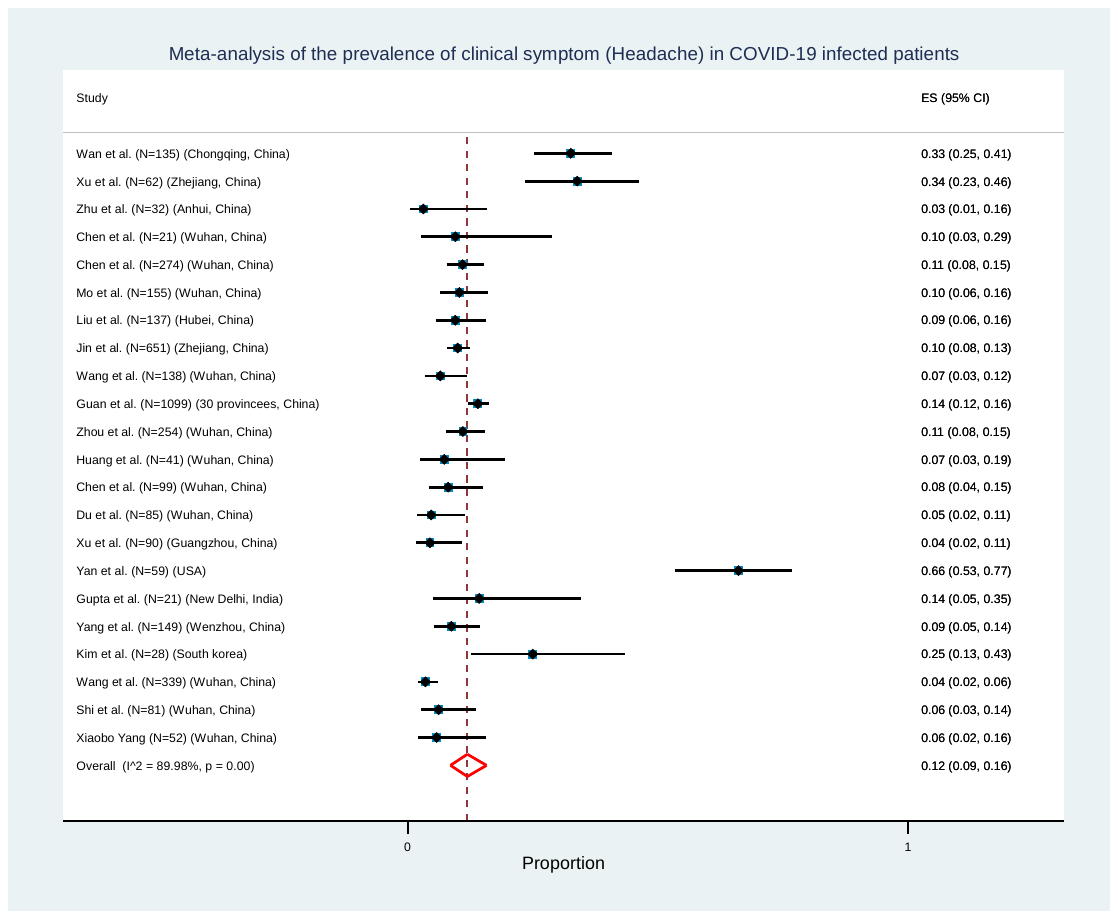

**S8. Diarrhea**

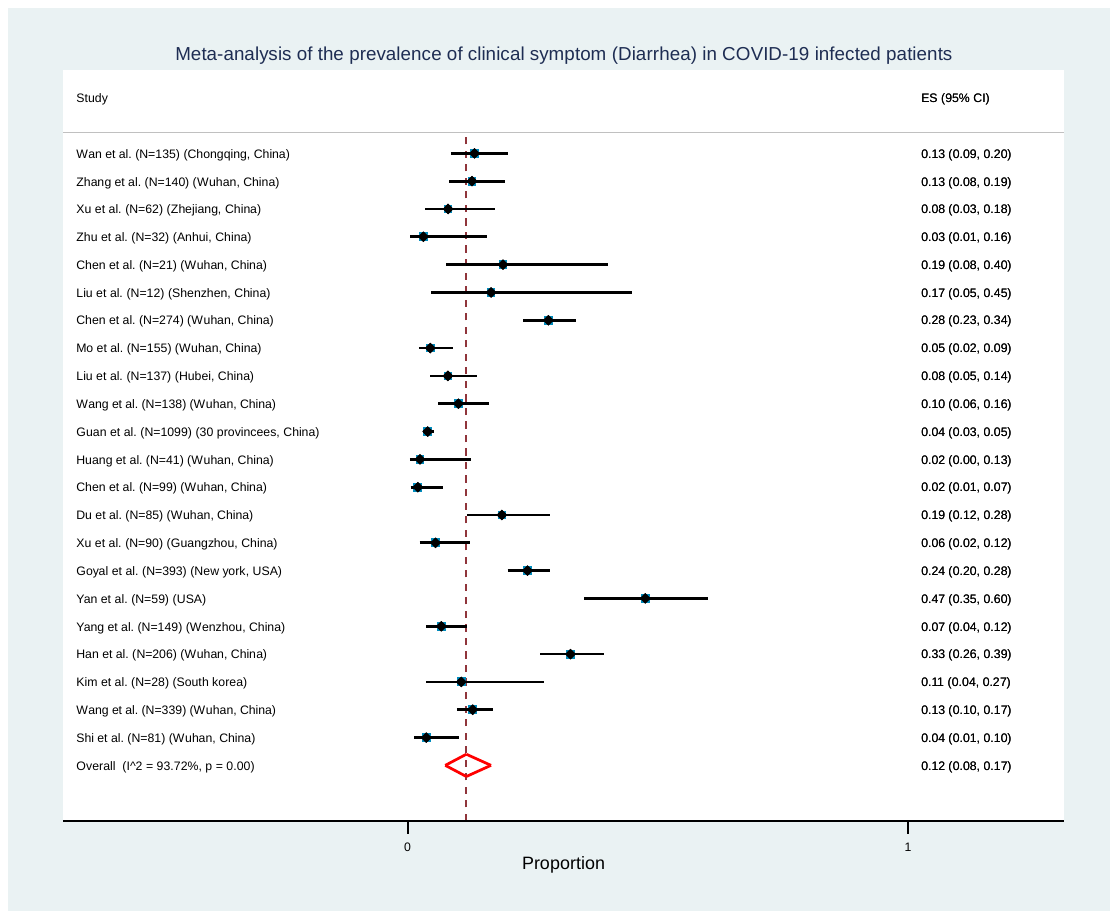

**S9. Sore Throat**

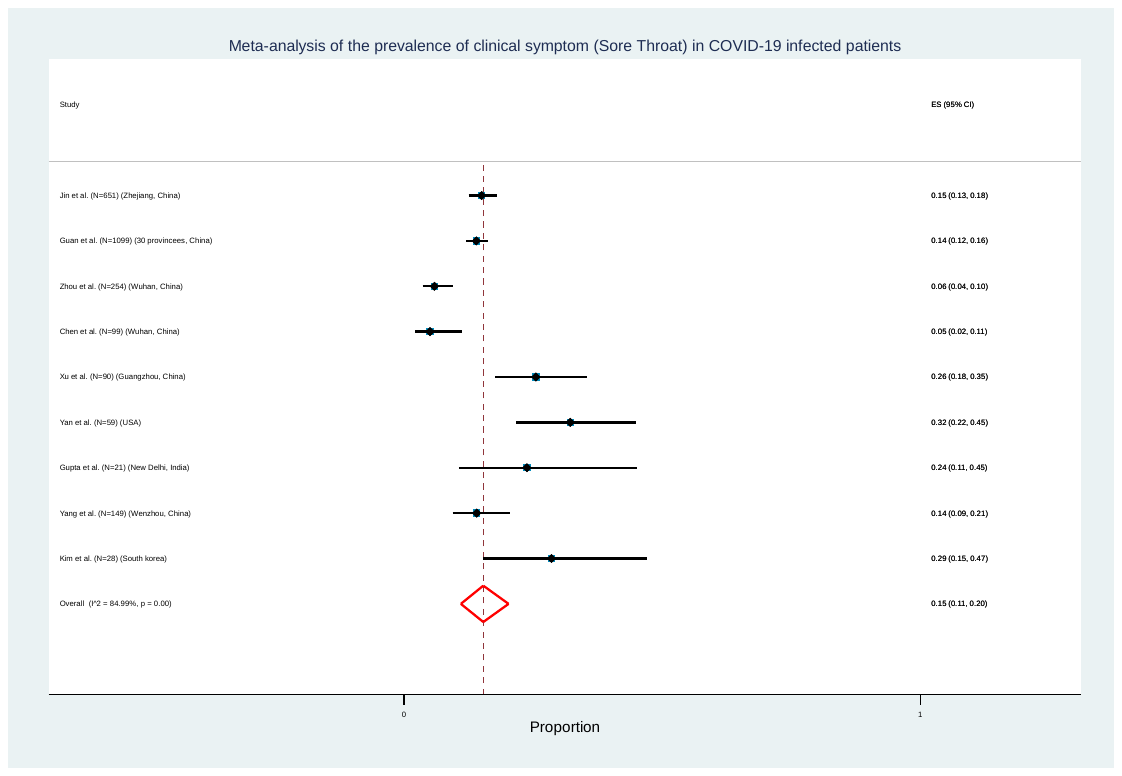

**S10. Myalgia/Muscle Ache**

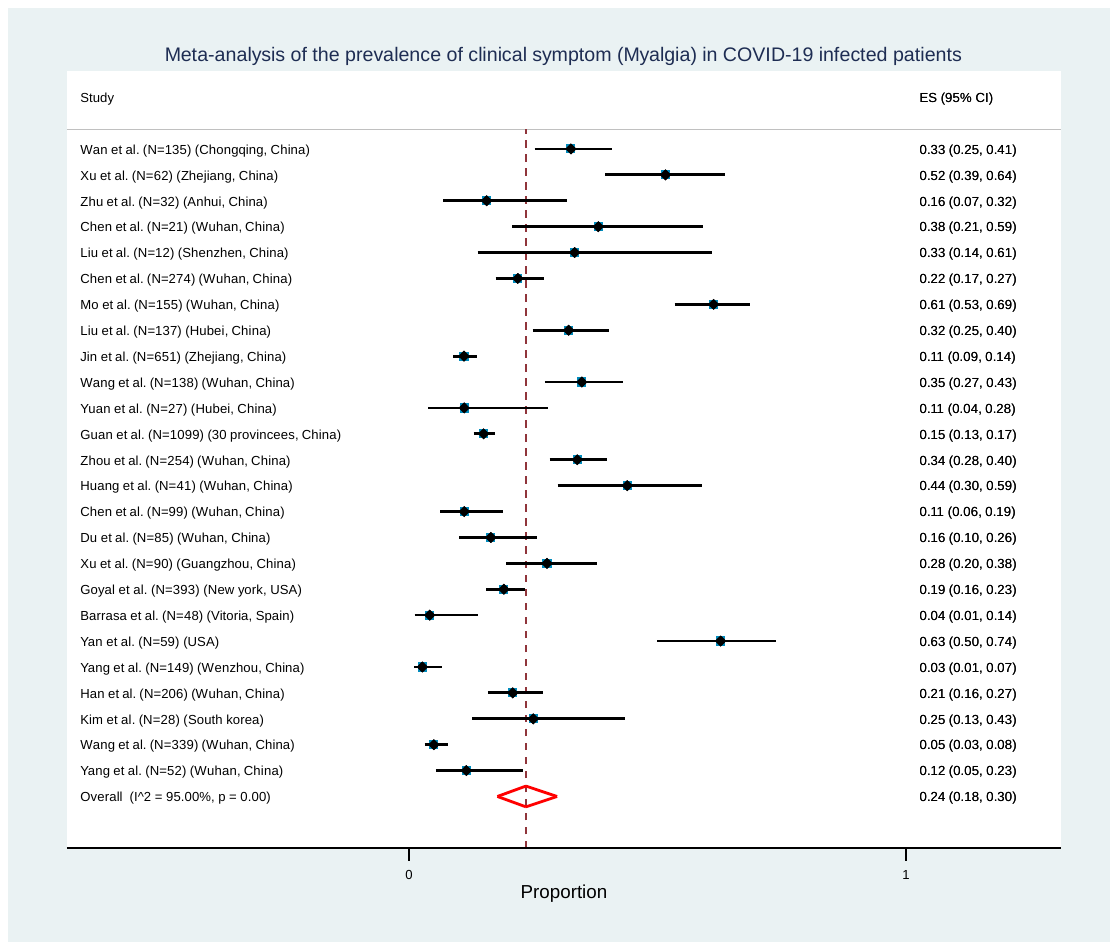

**S11. Rhinorrhea**

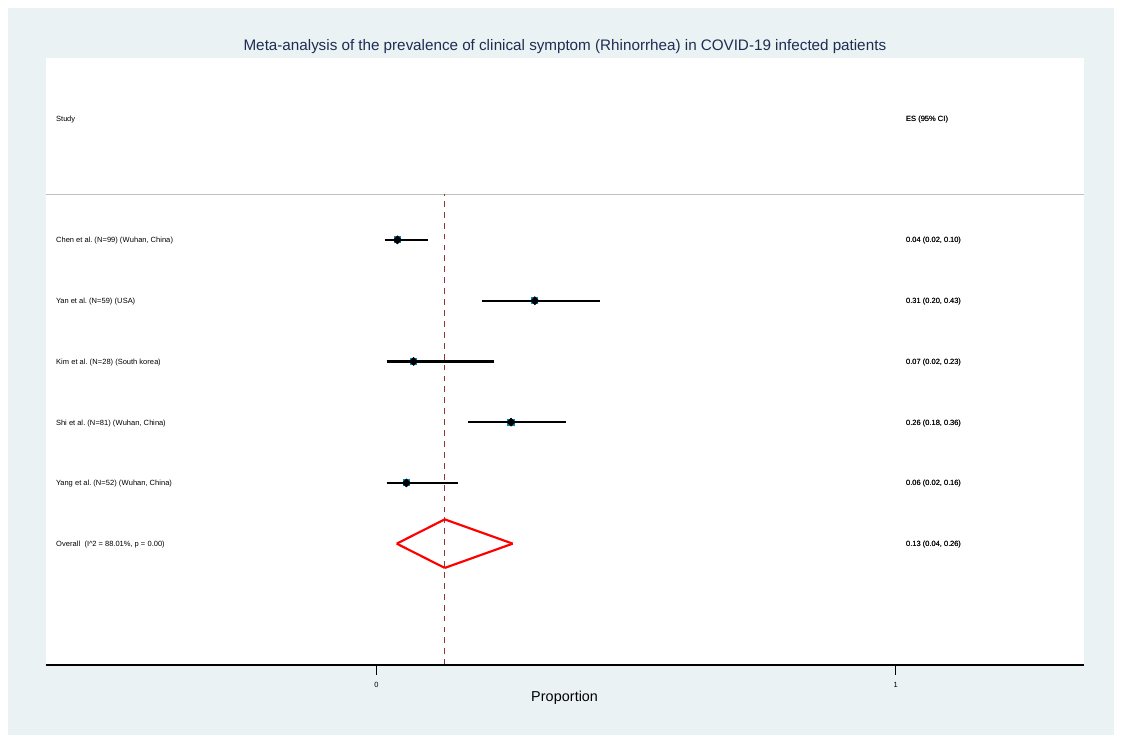

**S12. Sputum Production/Expectoration**

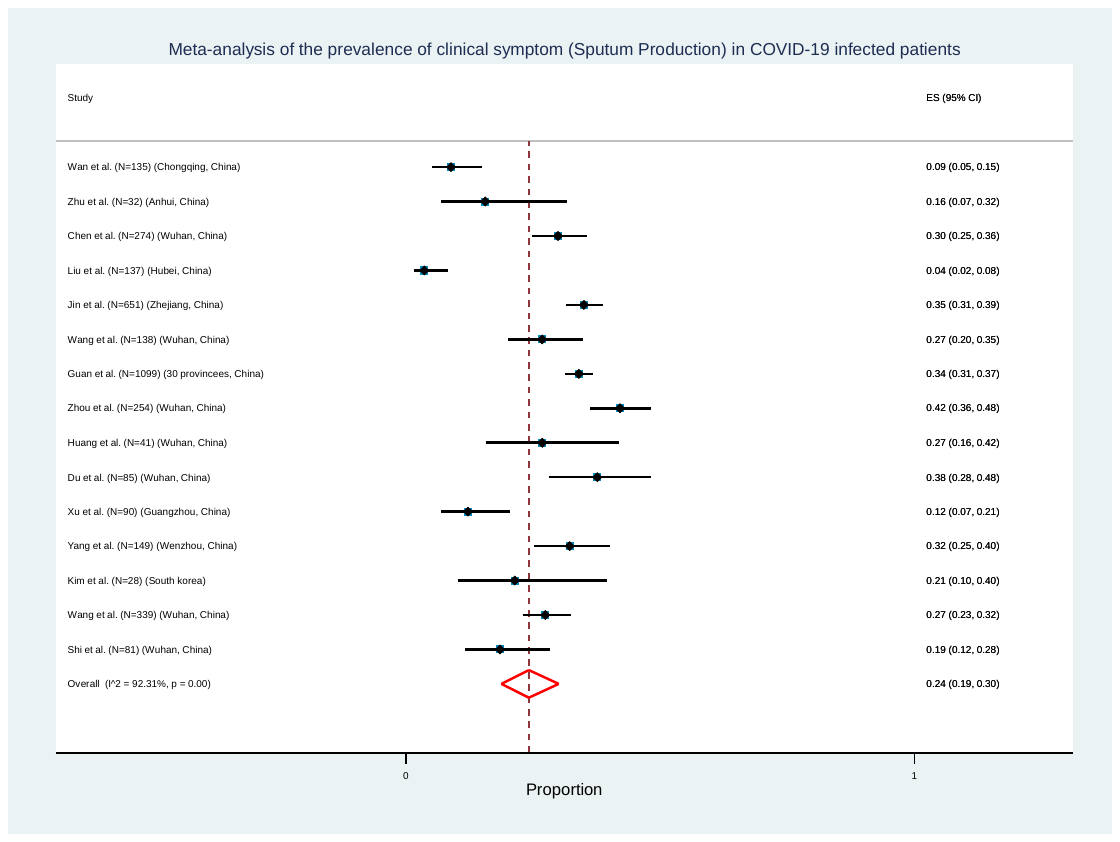

**S13. Chest tightness**

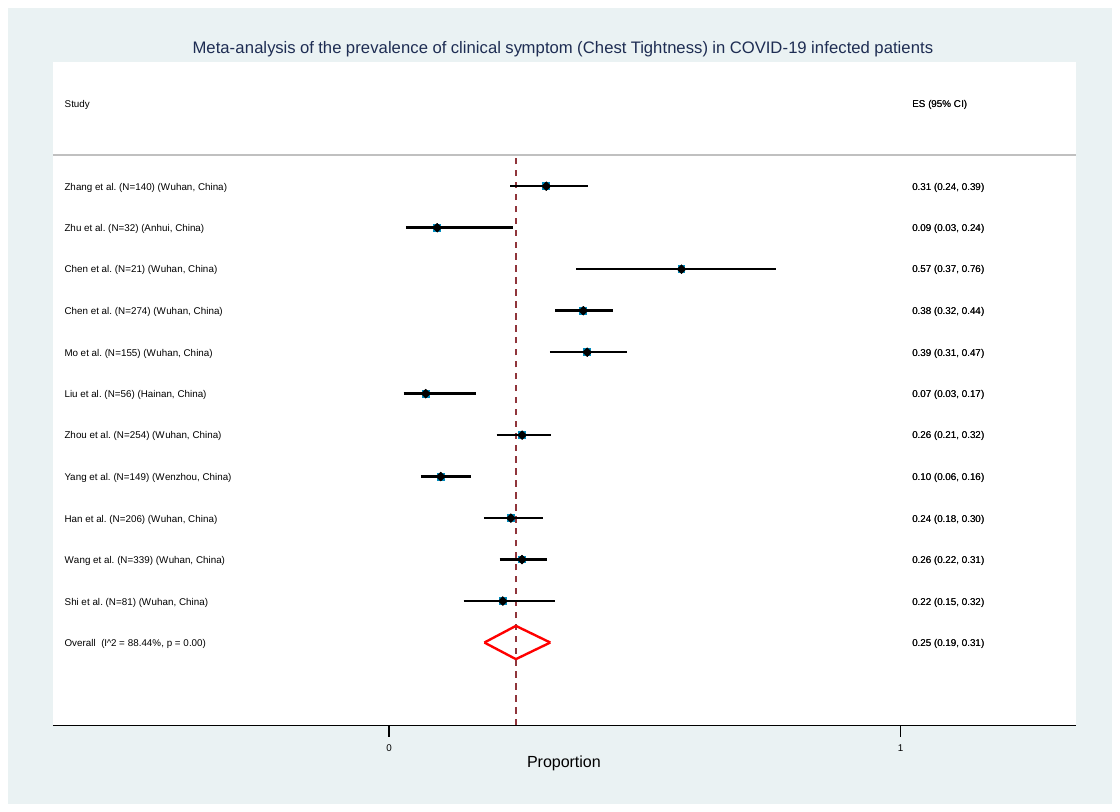

**S14. Chest pain**

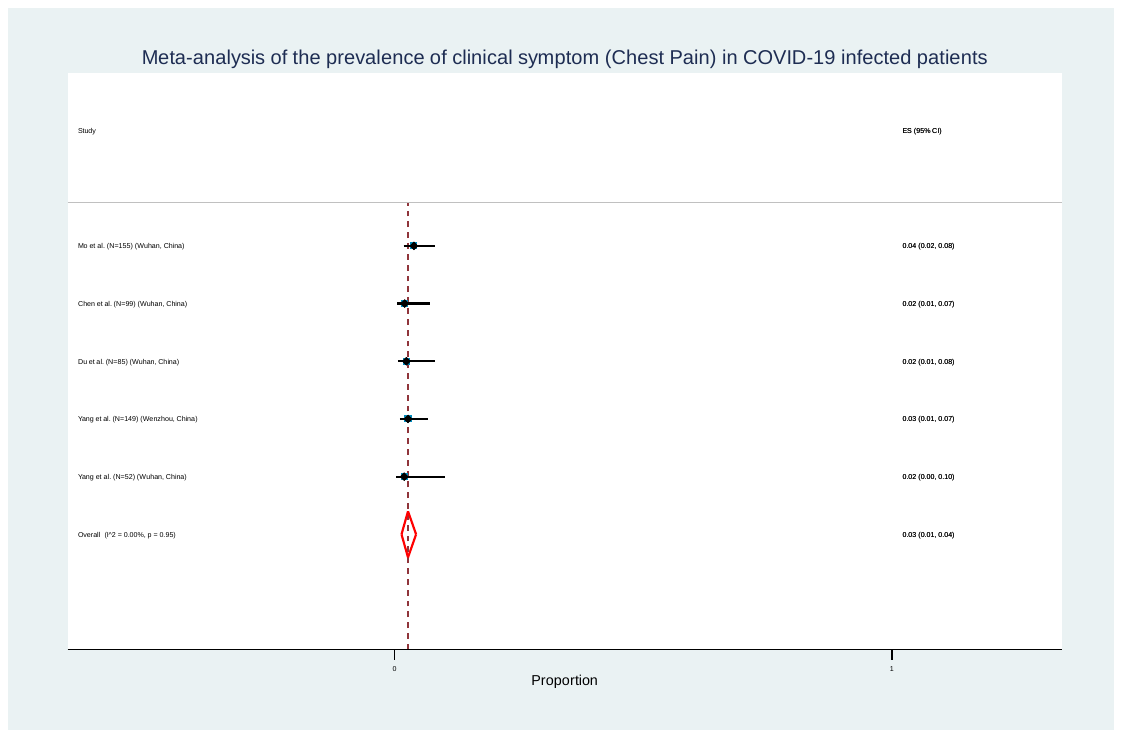

**S15. Nausea**

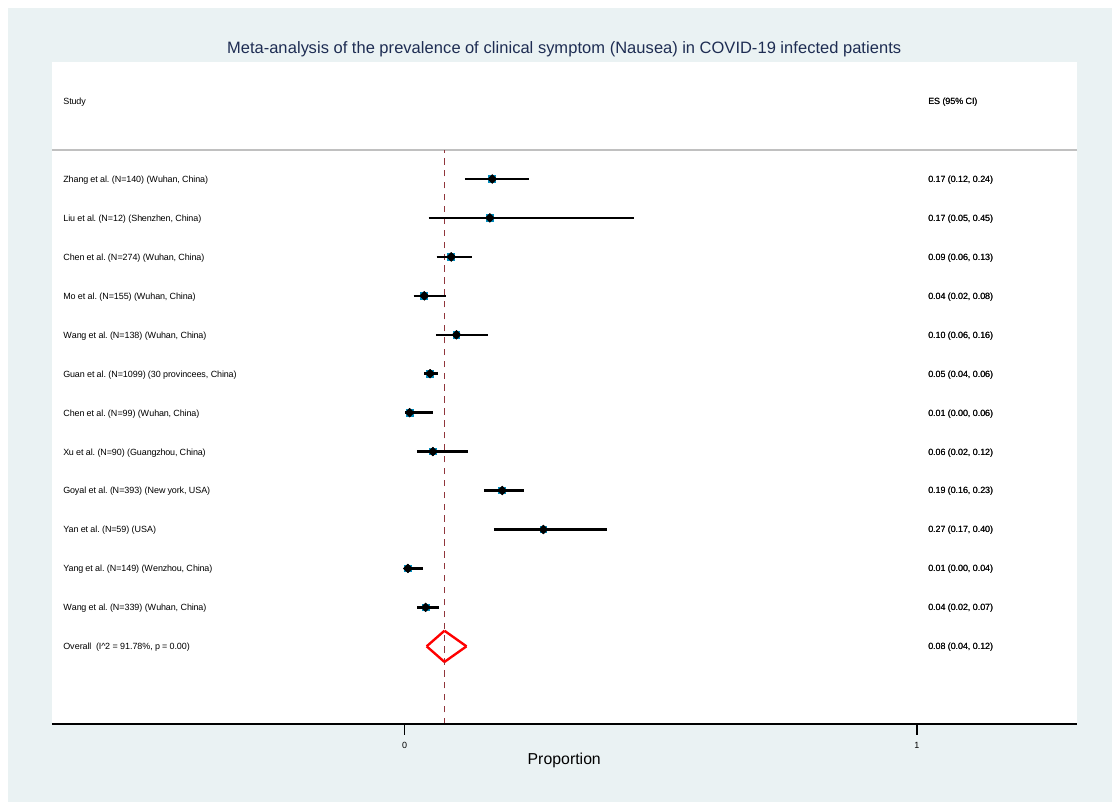

**S16. Vomiting**

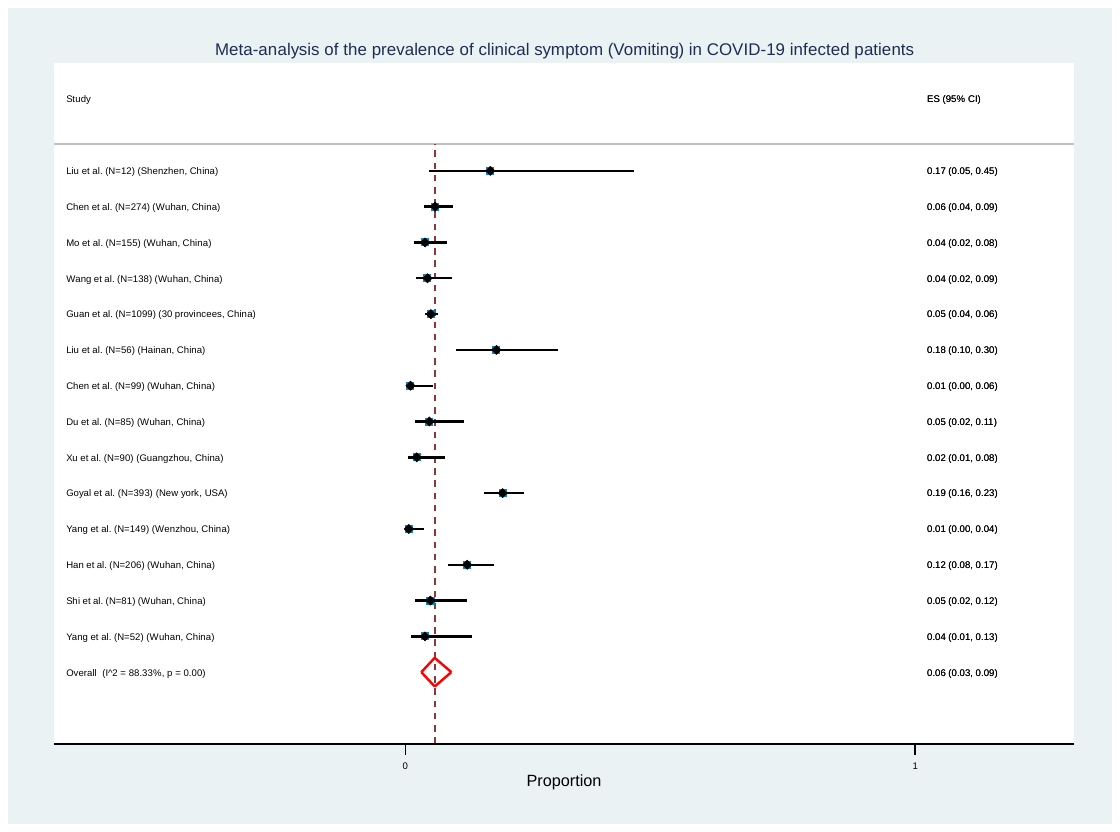

**S17. Abdominal Pain**

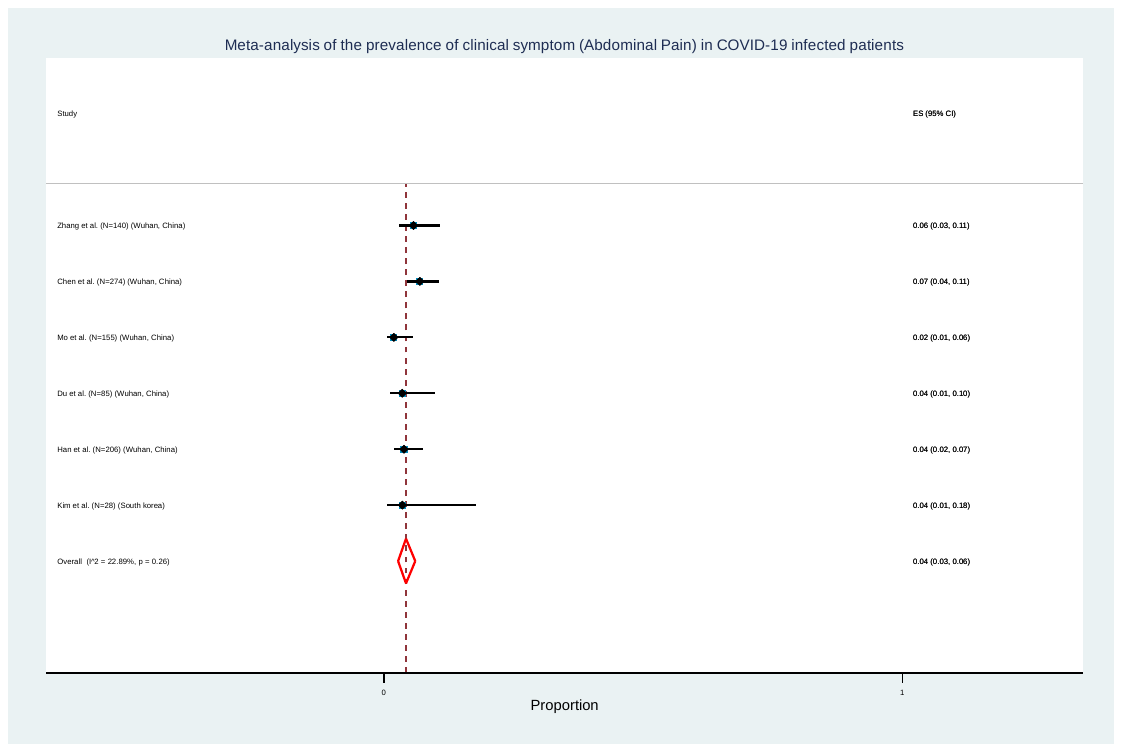

**S18. Dizziness**

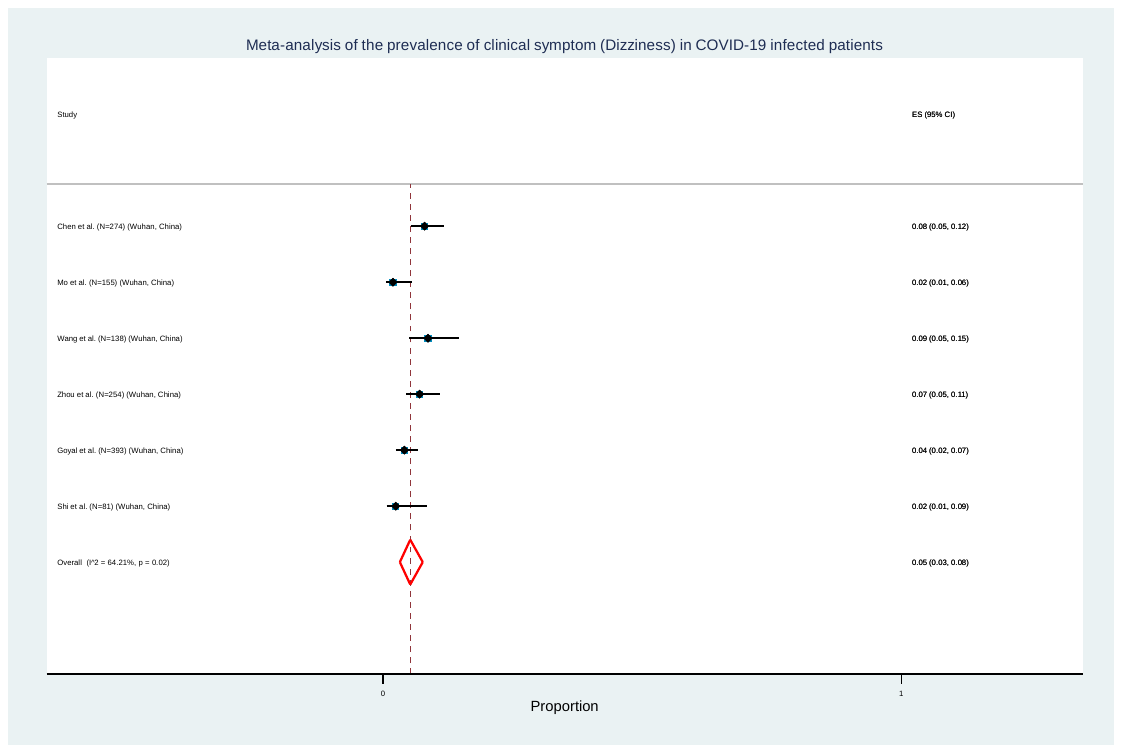

**S19. Anorexia**

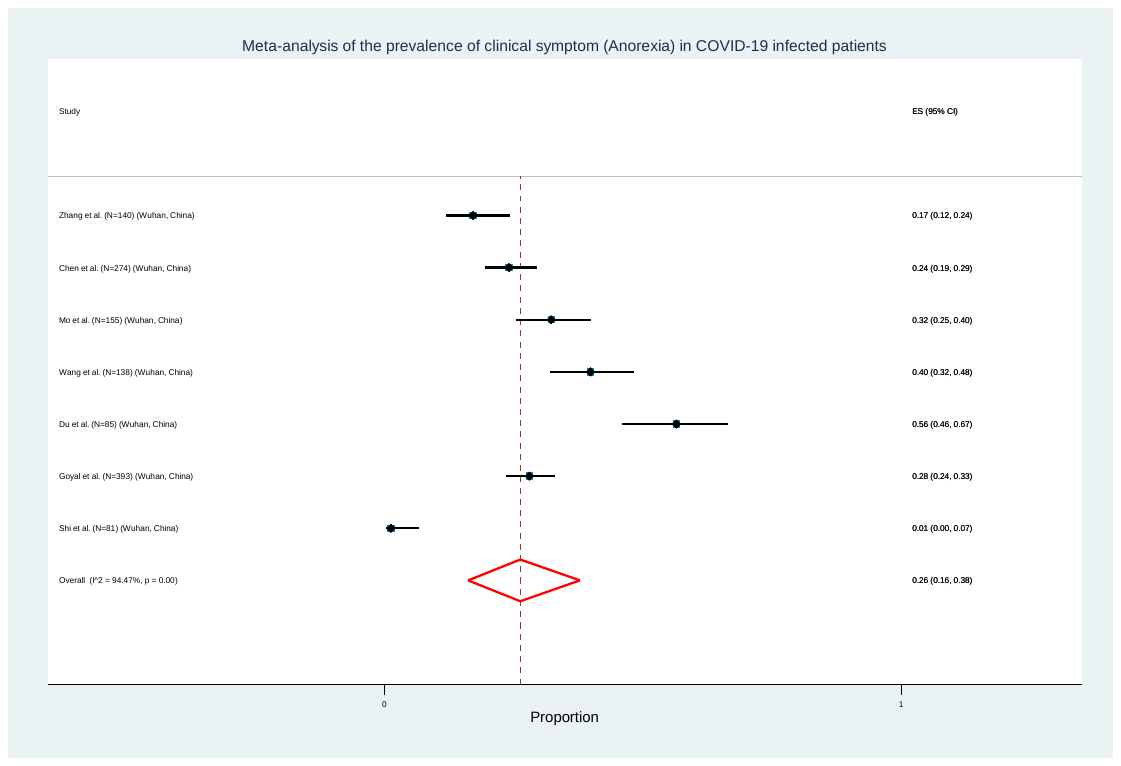

**S20. Pharyngalgia**

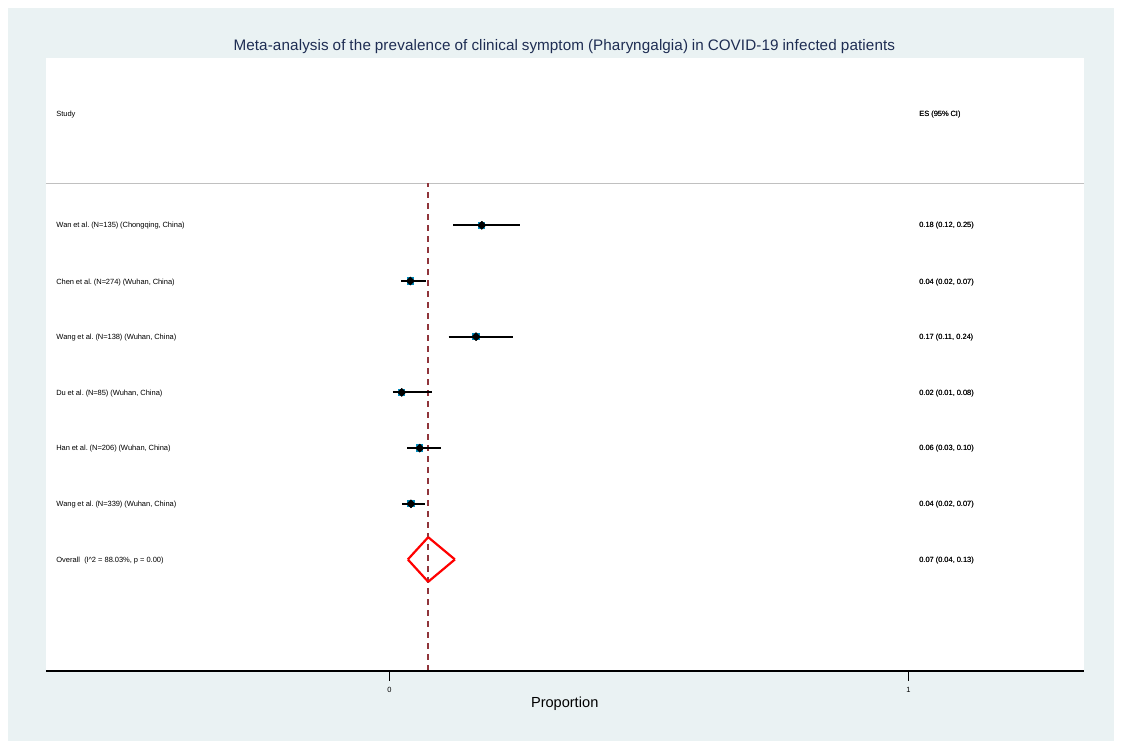

**S21. Haemoptysis**

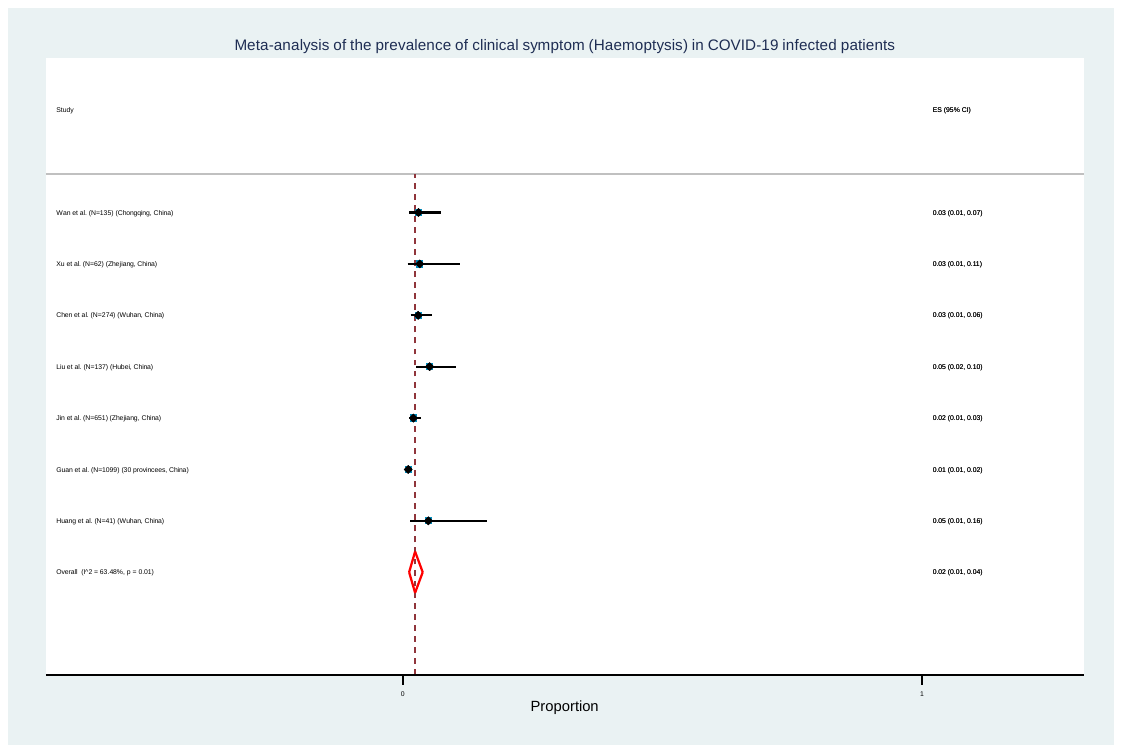

**Comorbidity**

**S22. Diabetes**

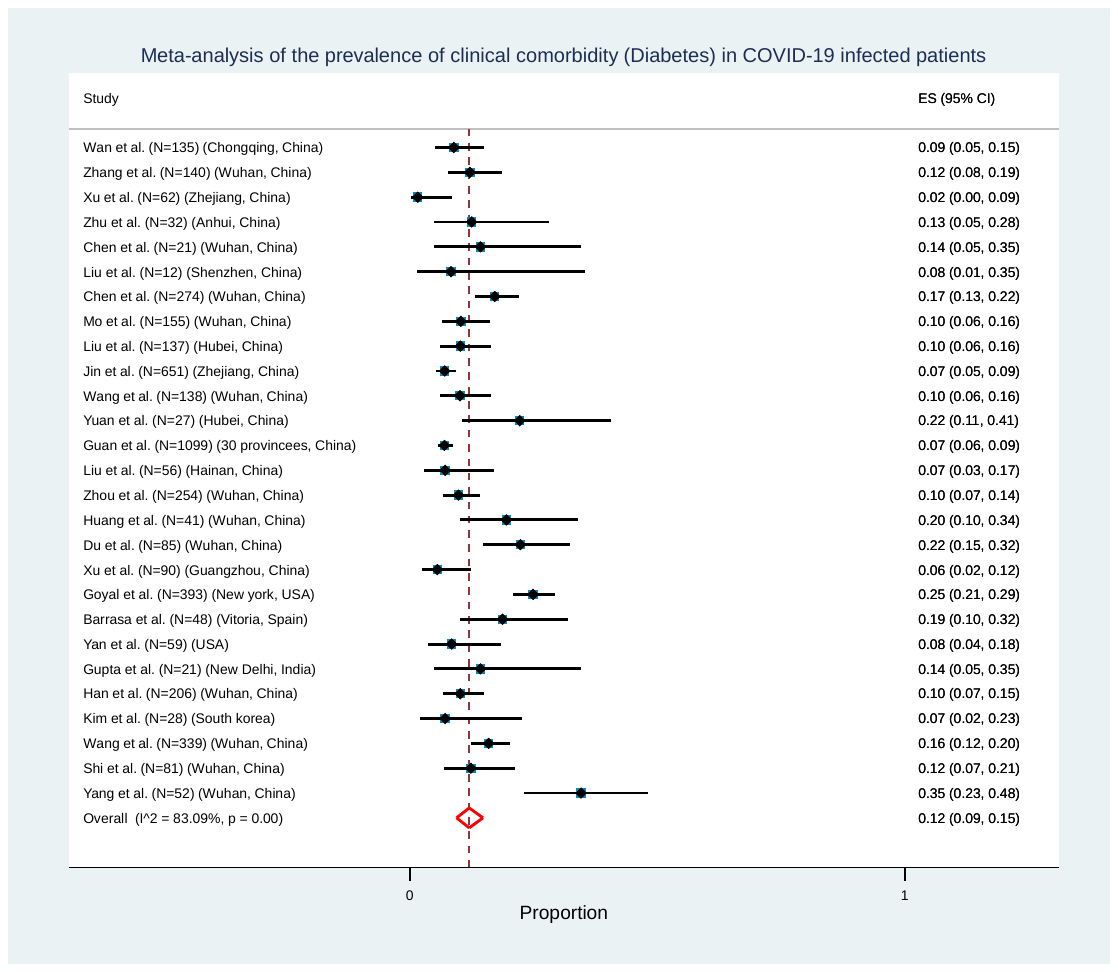

**S23. Hypertension**

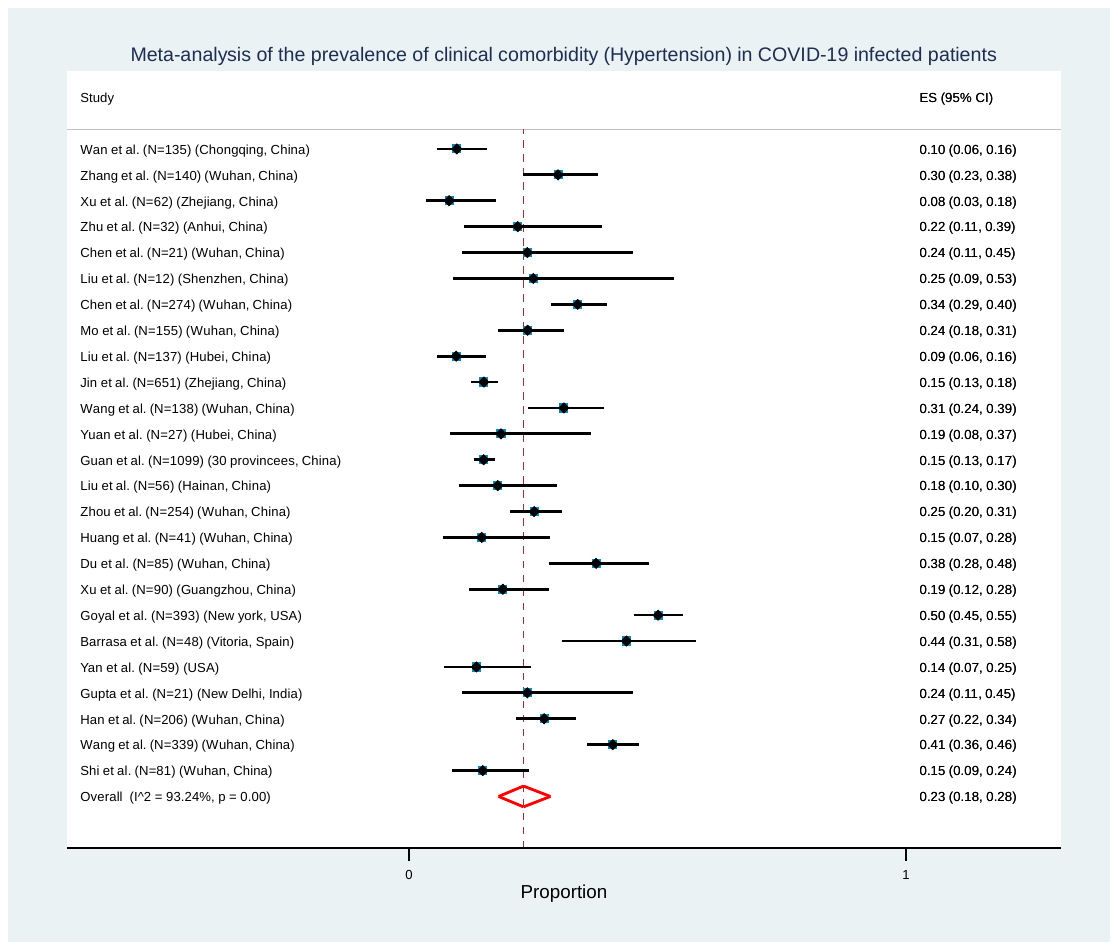

**S24. Cardiovascular Disease**

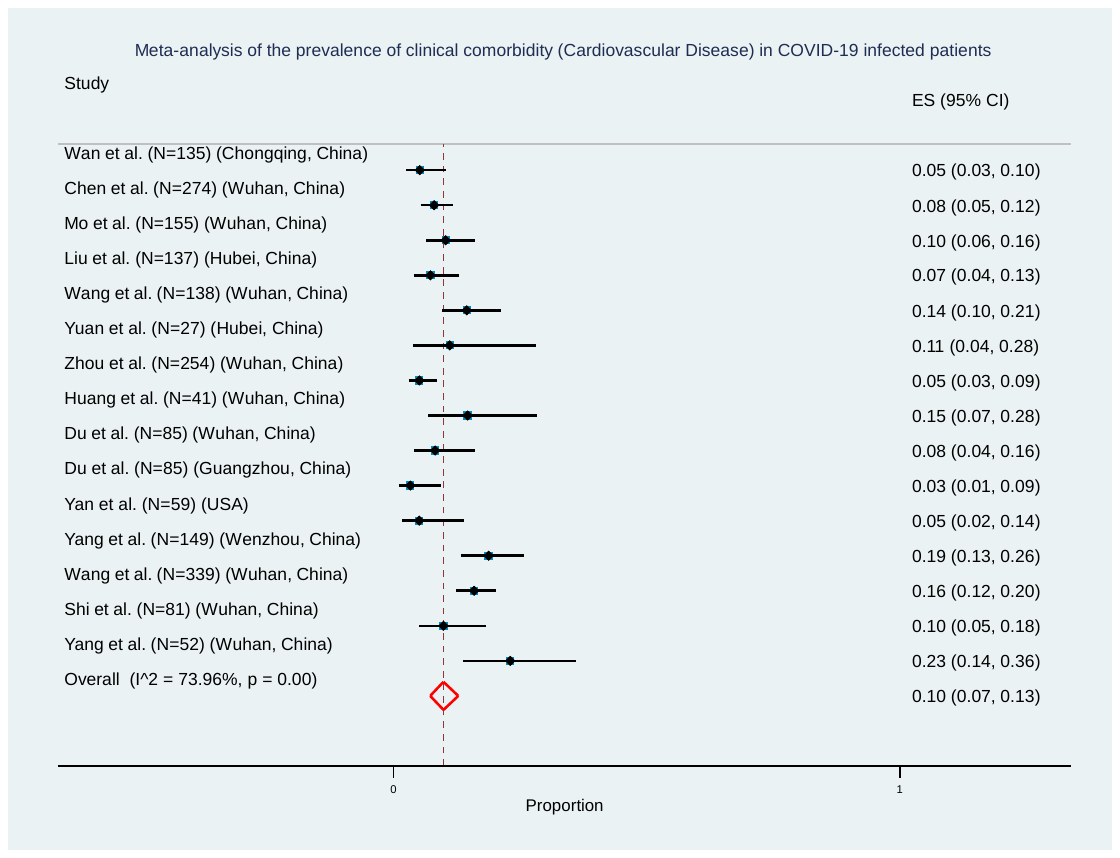

**S25. Coronary heart disease**

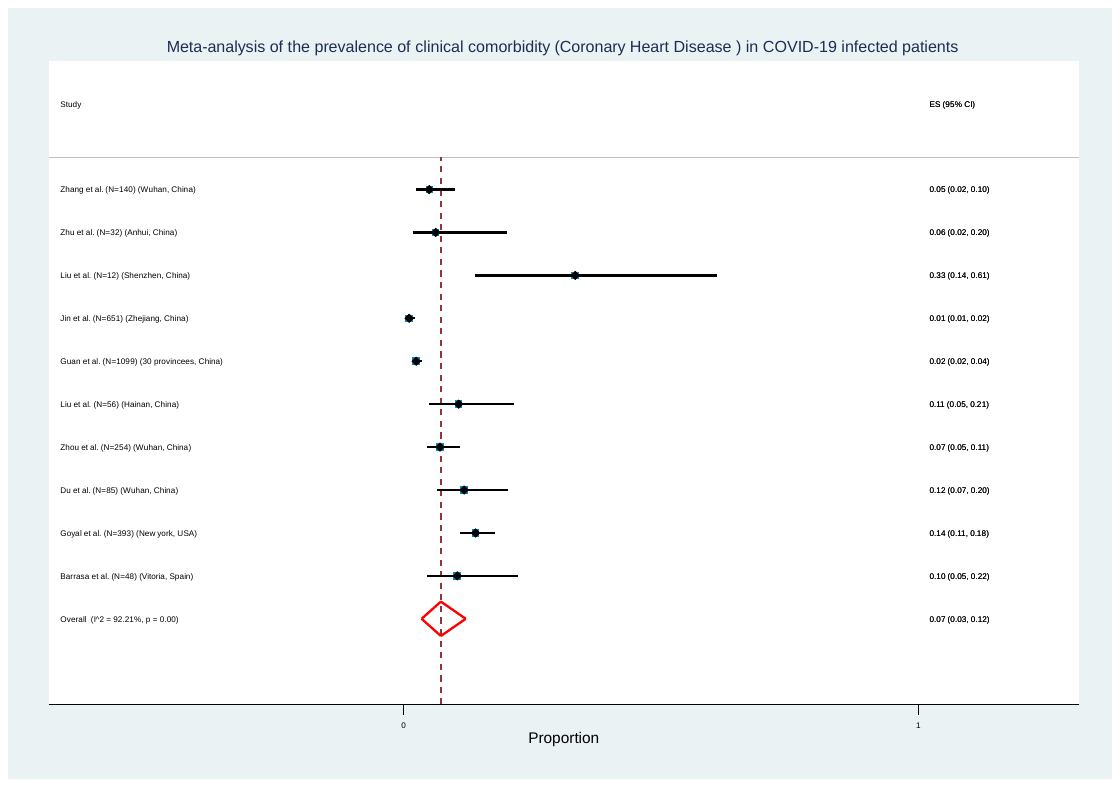

**S26. Cerebrovascular disease**

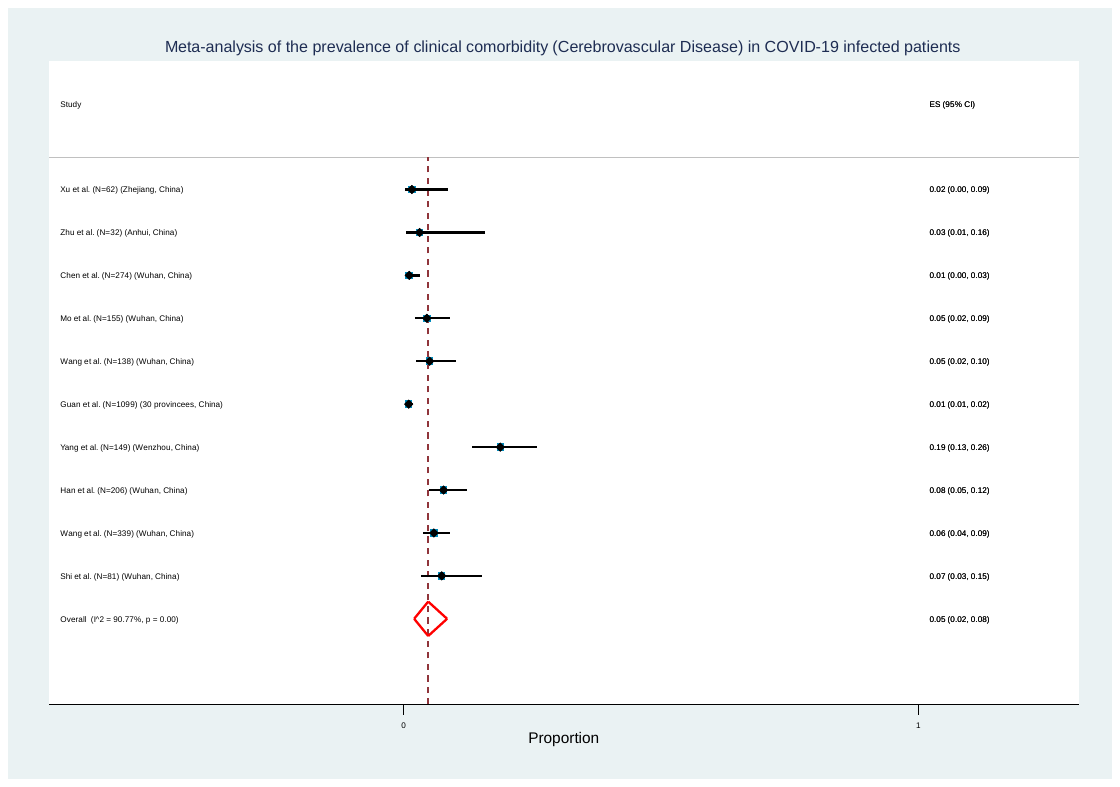

**S27. COPD/Lung disease**

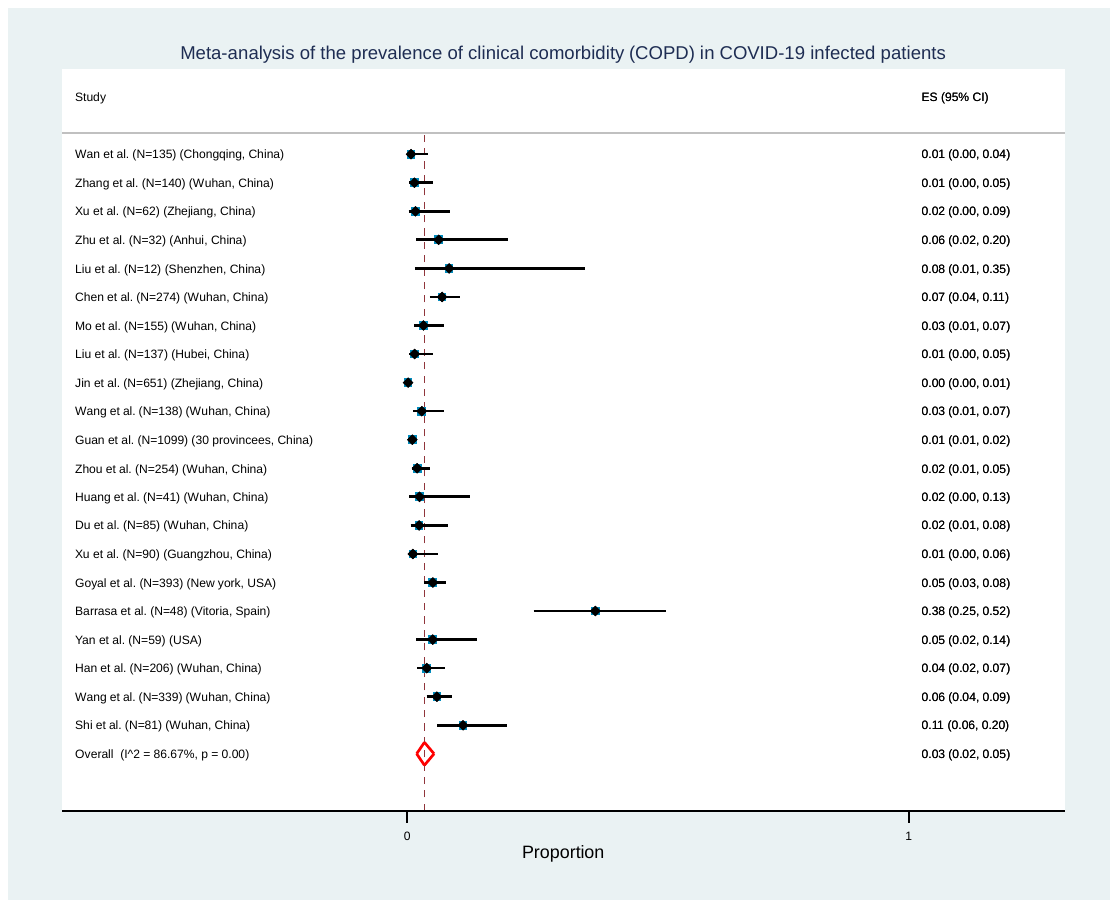

**S28. Chronic liver disease**

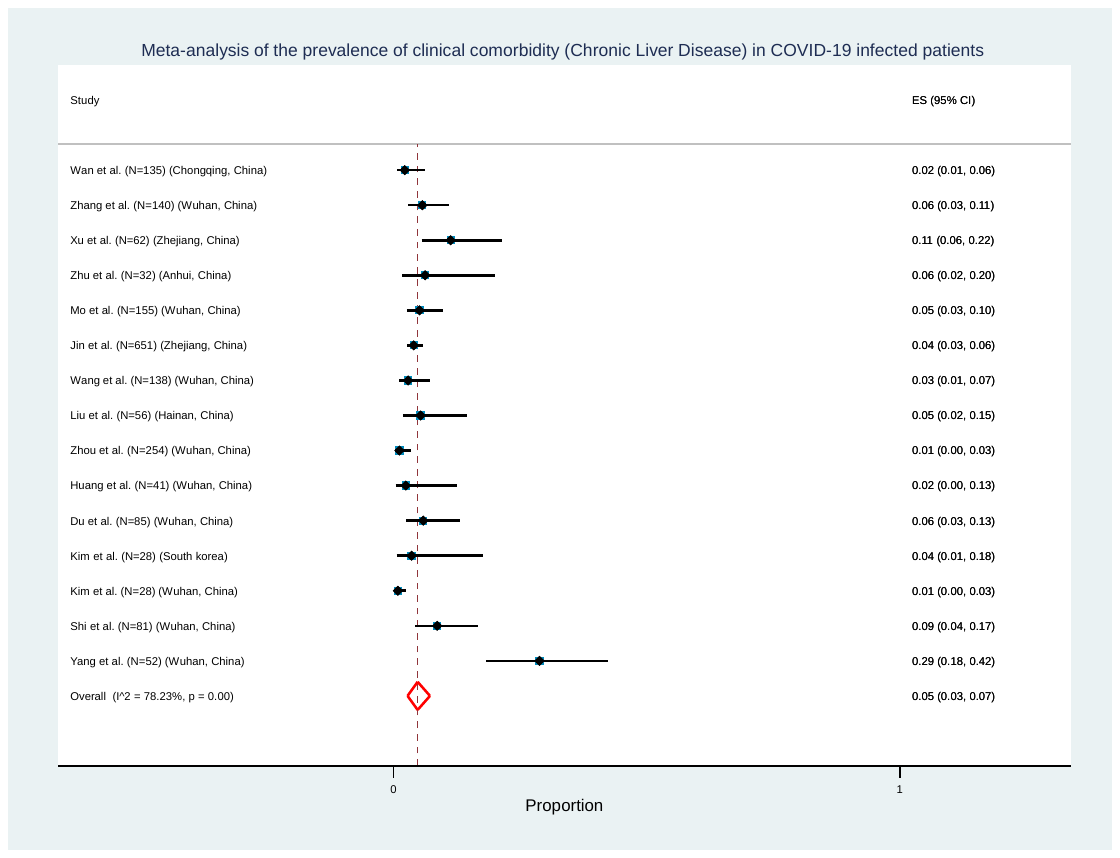

**S29. Chronic Renal disease**

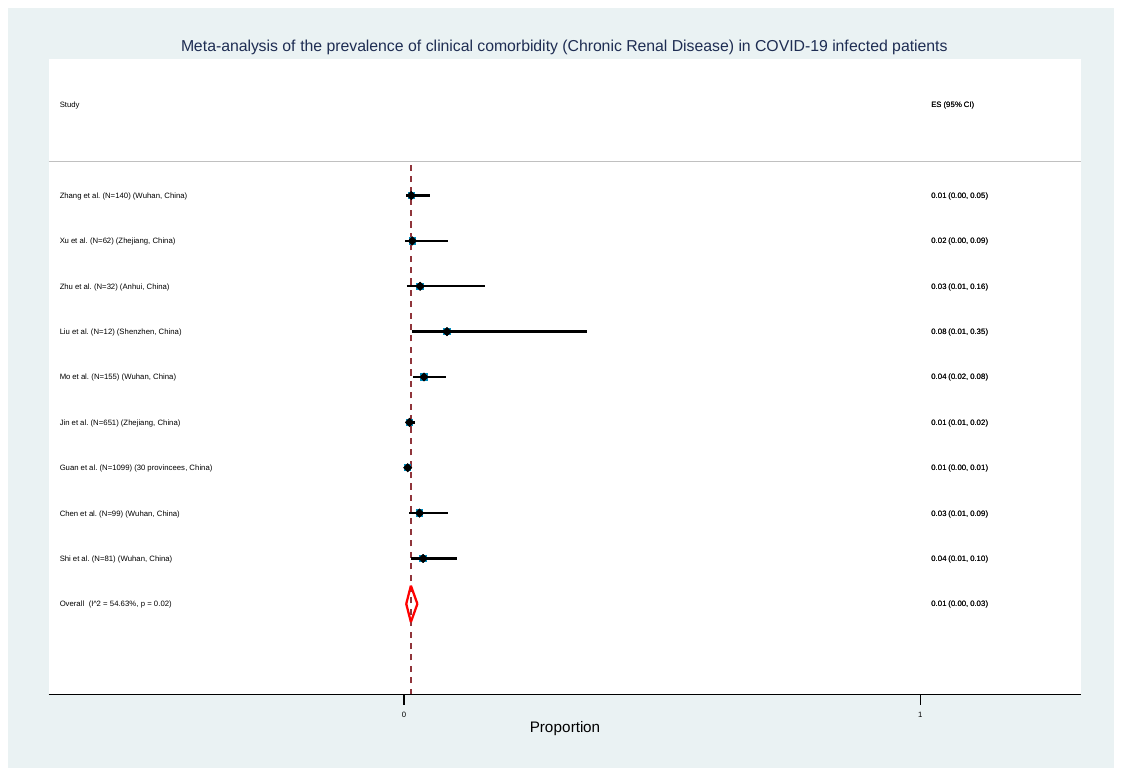

**S30. Chronic Kidney disease**

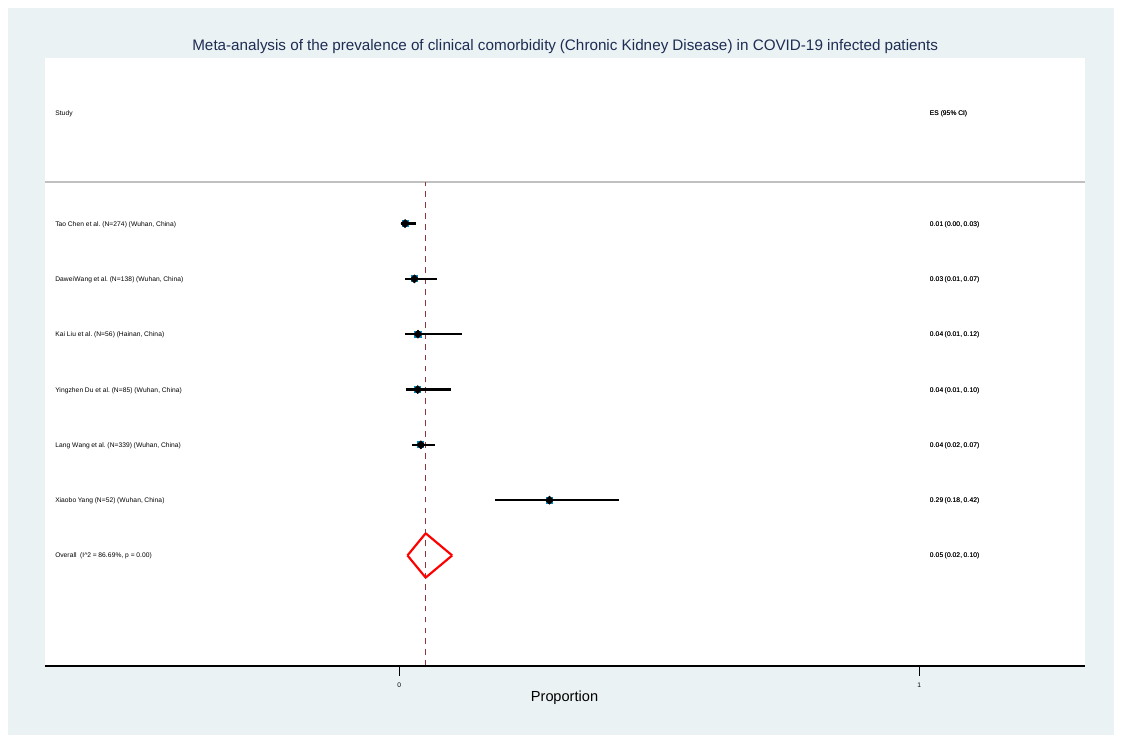

**S31. Malignancy**

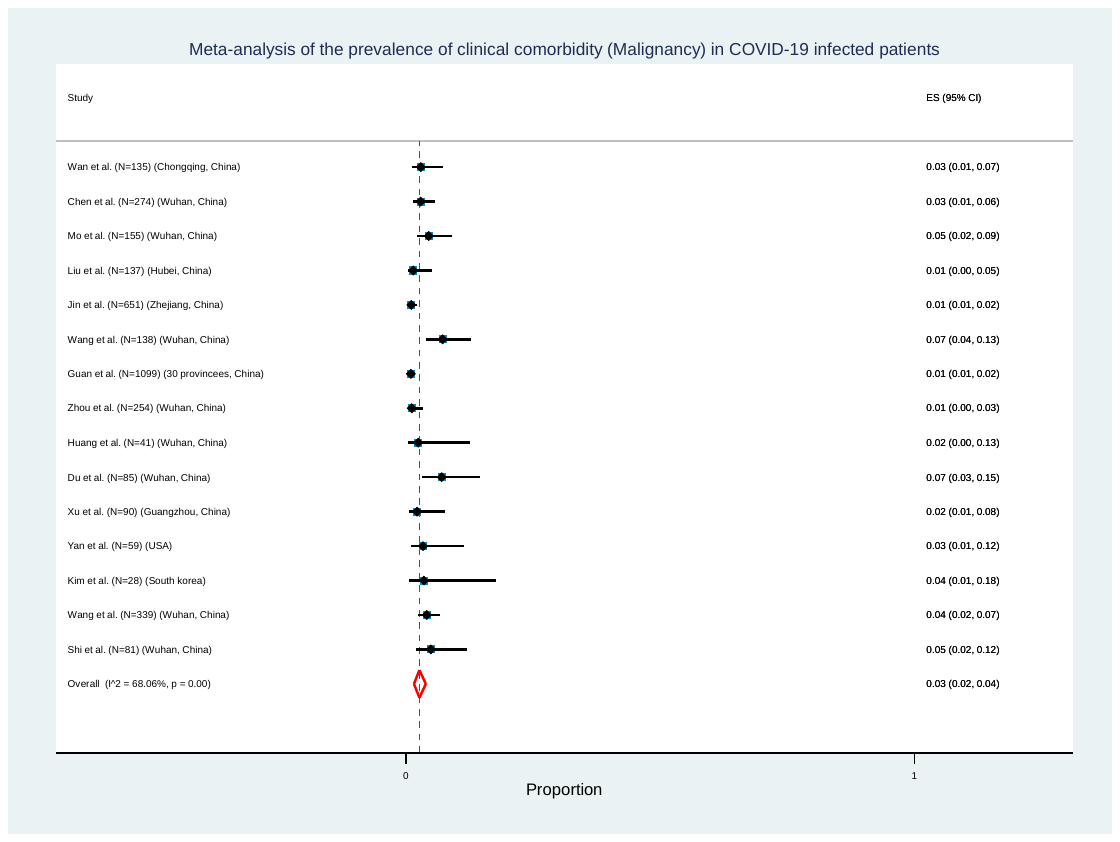

**S32. ARDS**

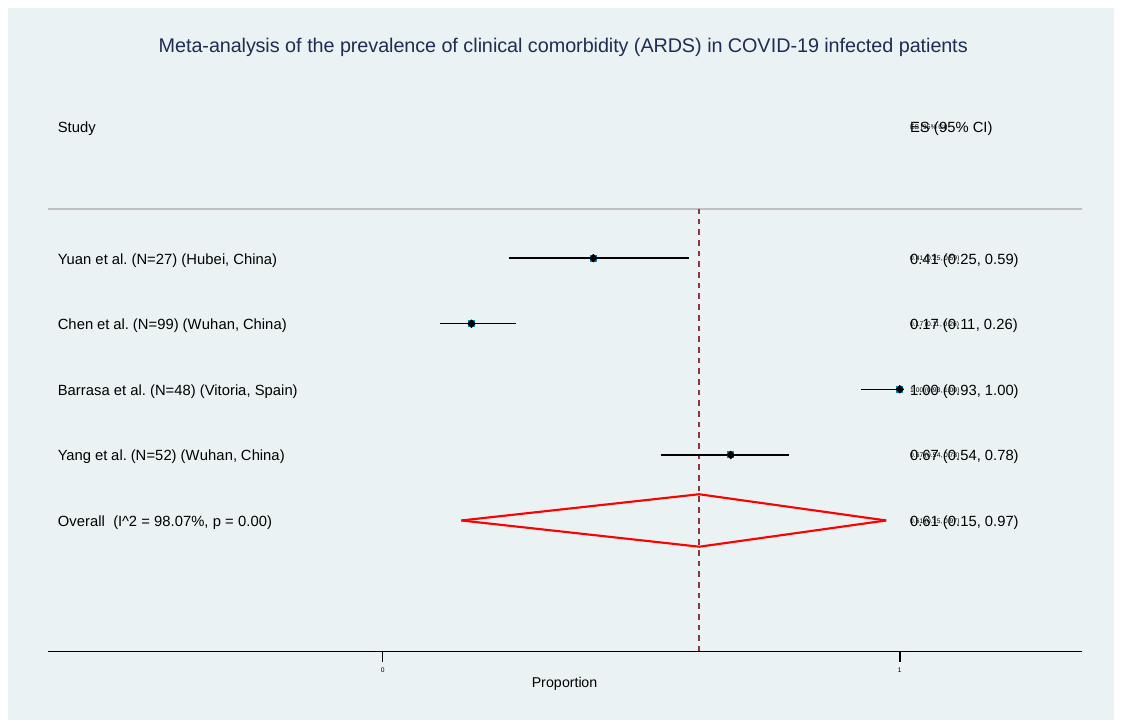

**Funnel Plots of the symptoms and comorbidities**

**Symptoms**

**S33. Fever**

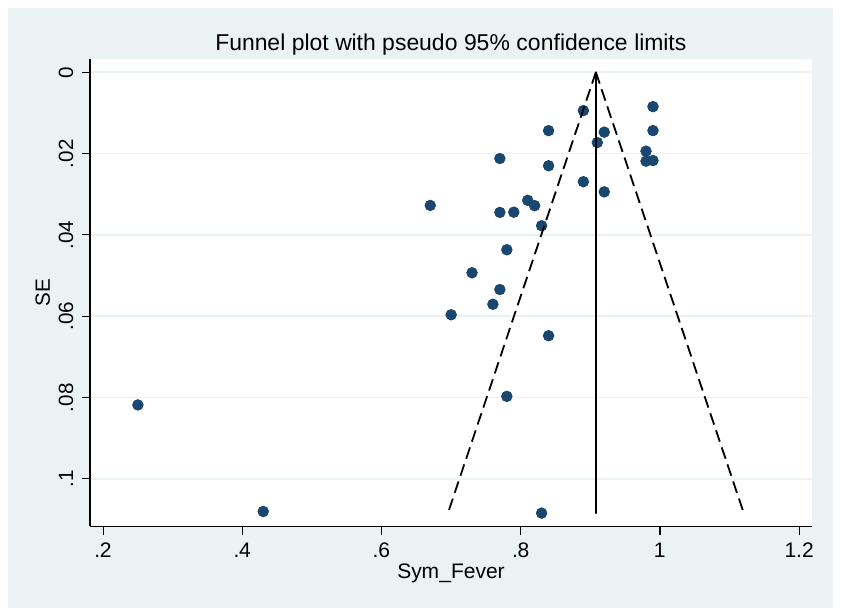

**S34. Cough / Dry cough**

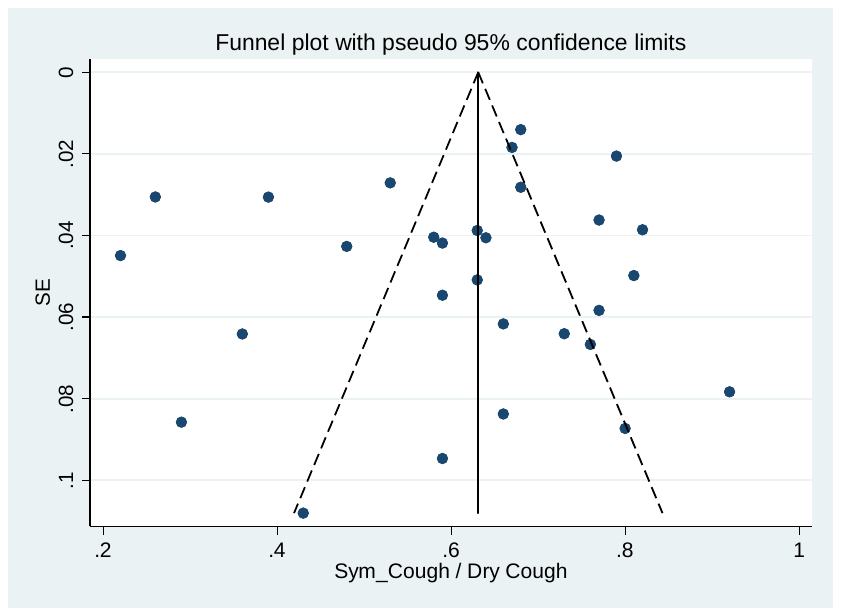

**S35. Fatigue/ Weakness**

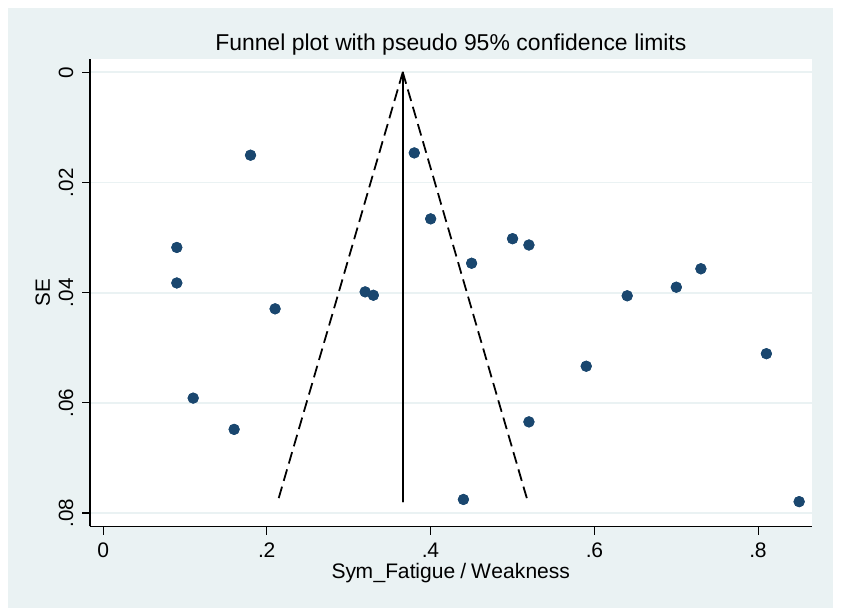

**S36. Dyspnea/ Shortness of breath**

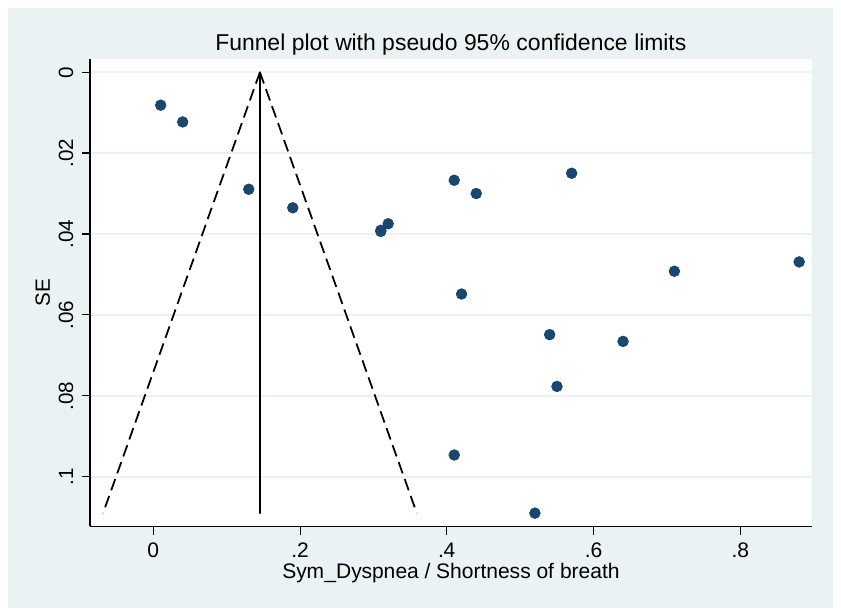

**S37. Headache**

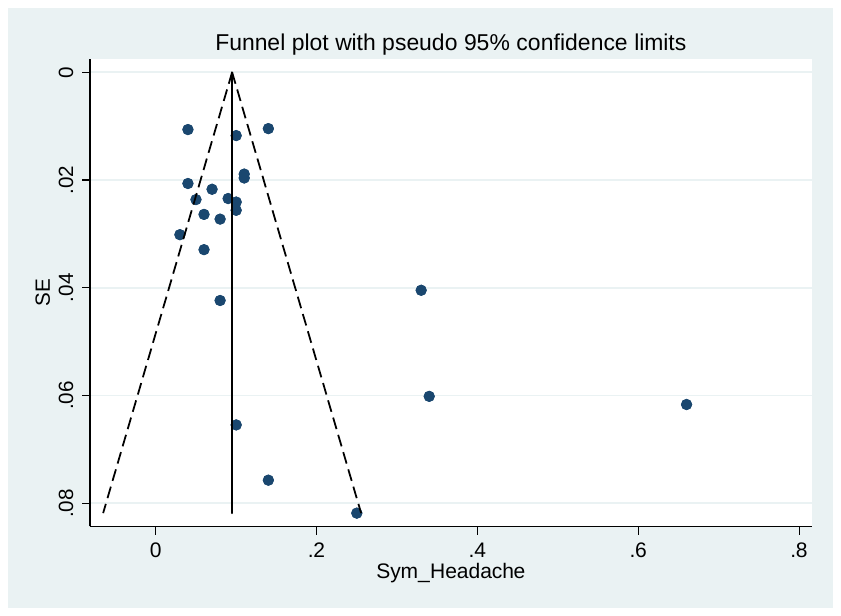

**S38. Diarrhea**

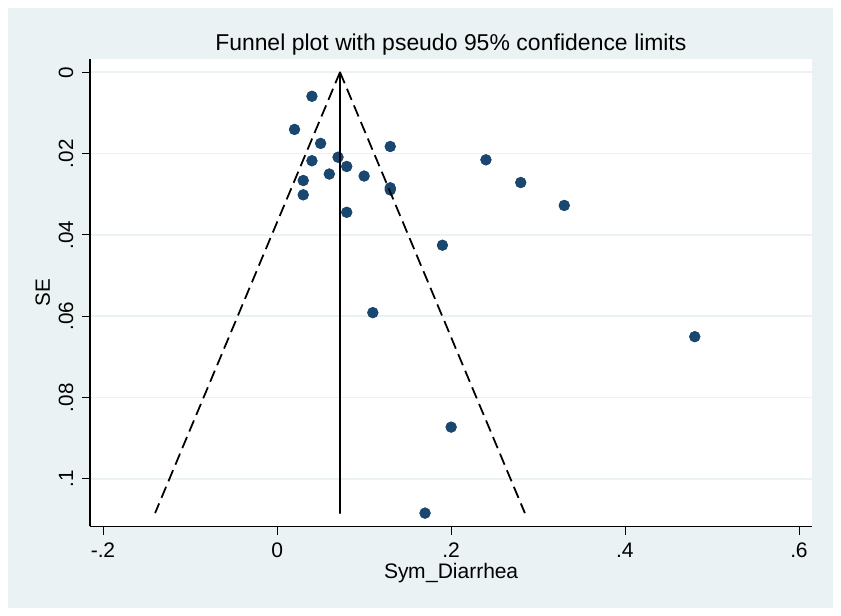

**S39. Sore Throat**

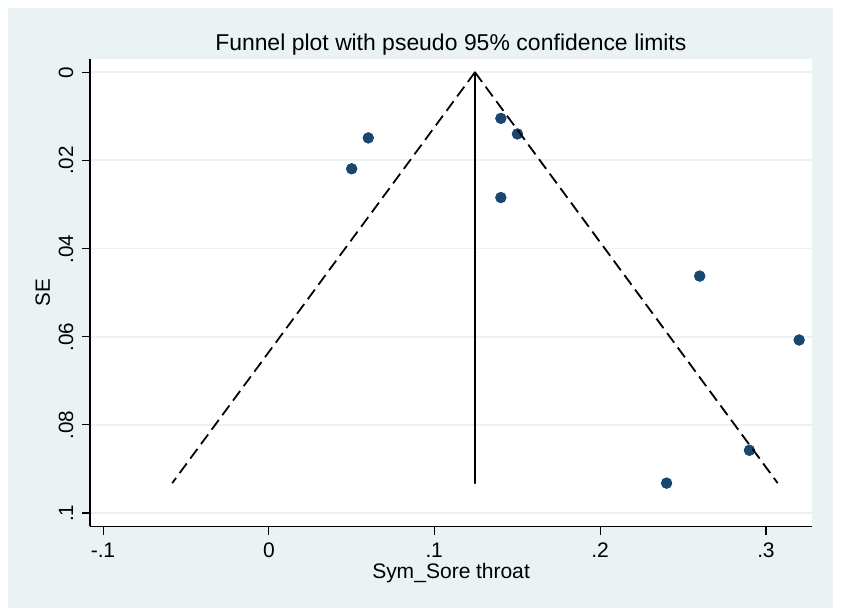

**S40. Myalgia / Muscle Ache**

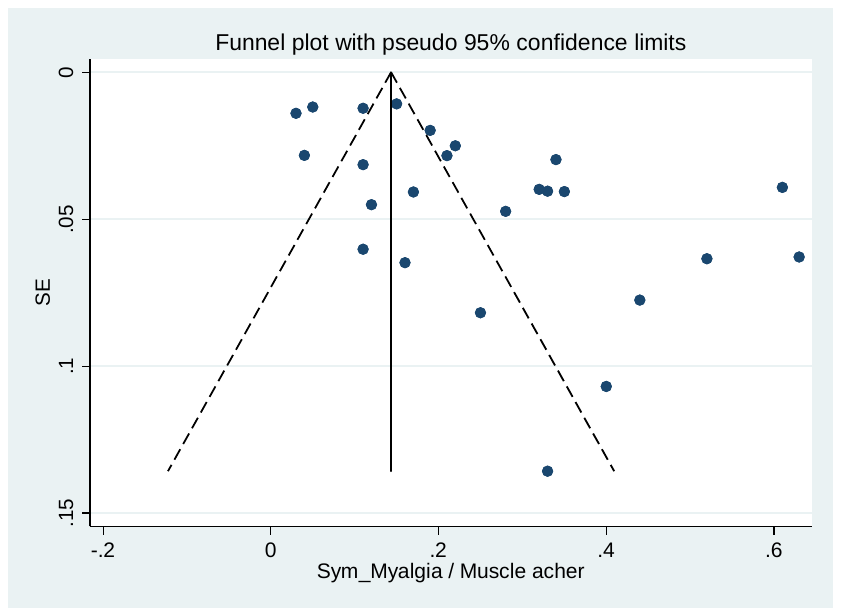

**S41. Rhinorrhea**

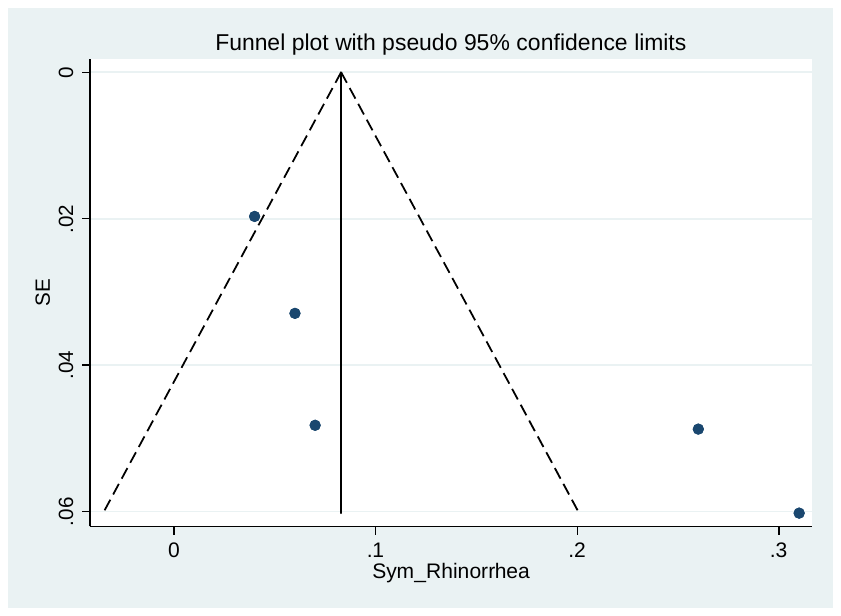

**S42. Sputum Production/Expectoration**

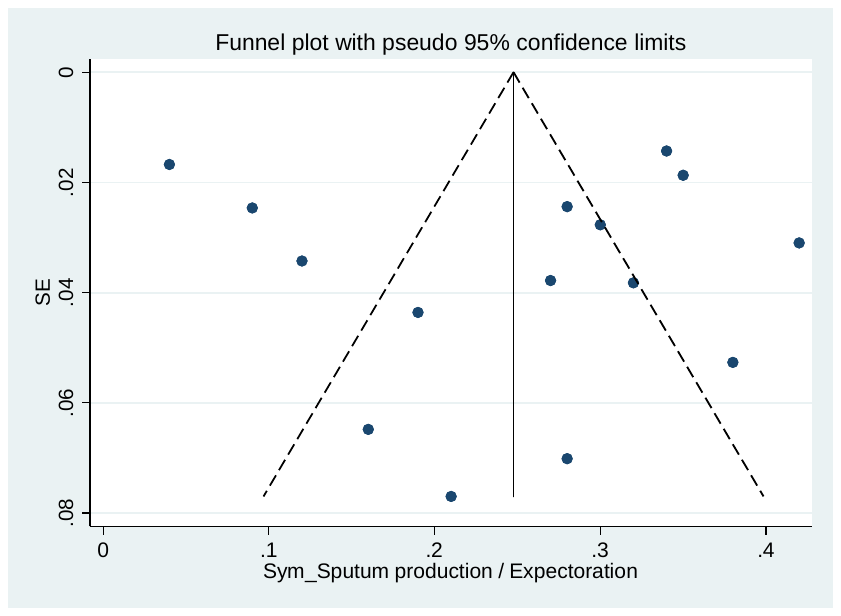

**S43. Chest Tightness**

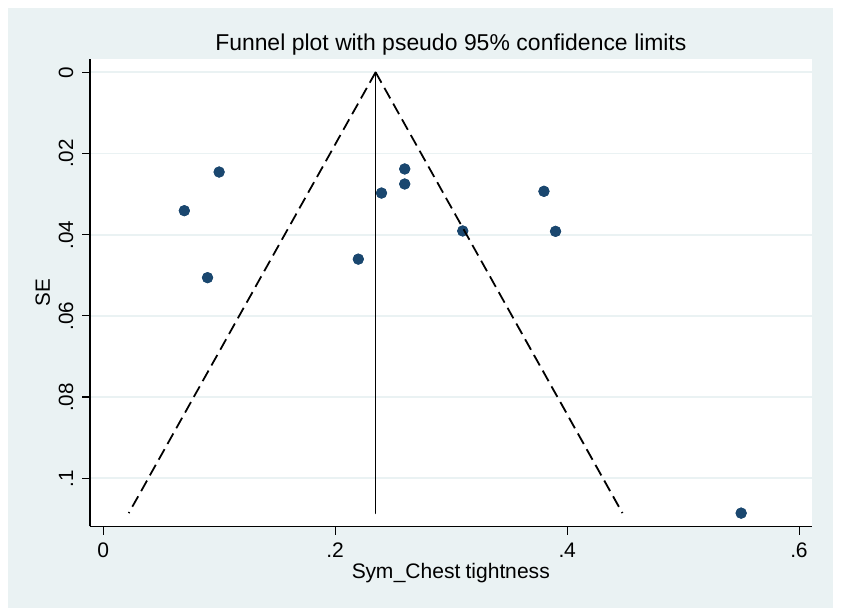

**S44. Chest Pain**

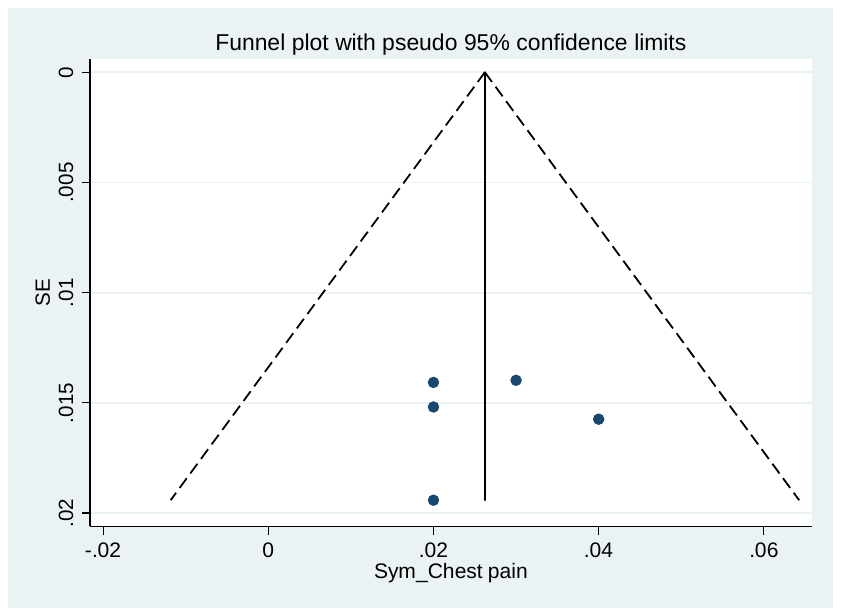

**S45. Nausea**

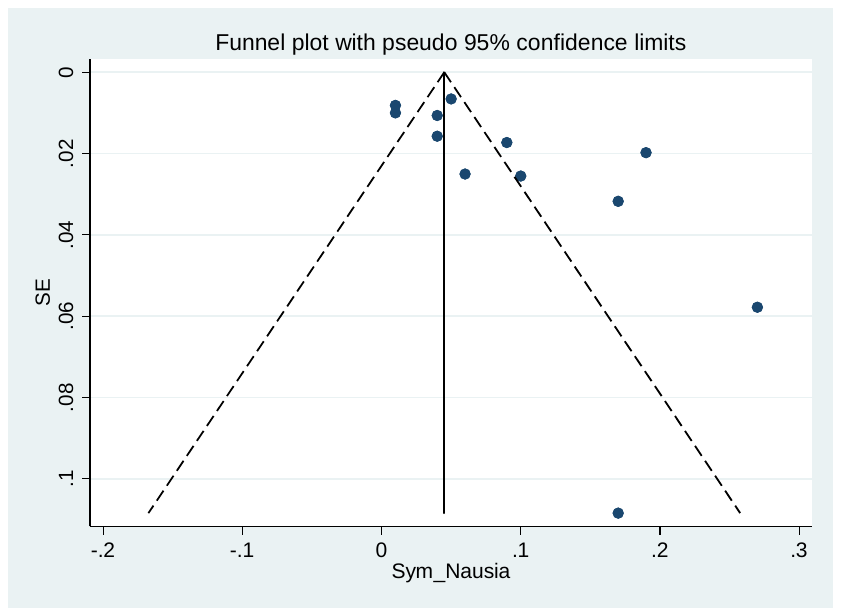

**S46.Vomiting**

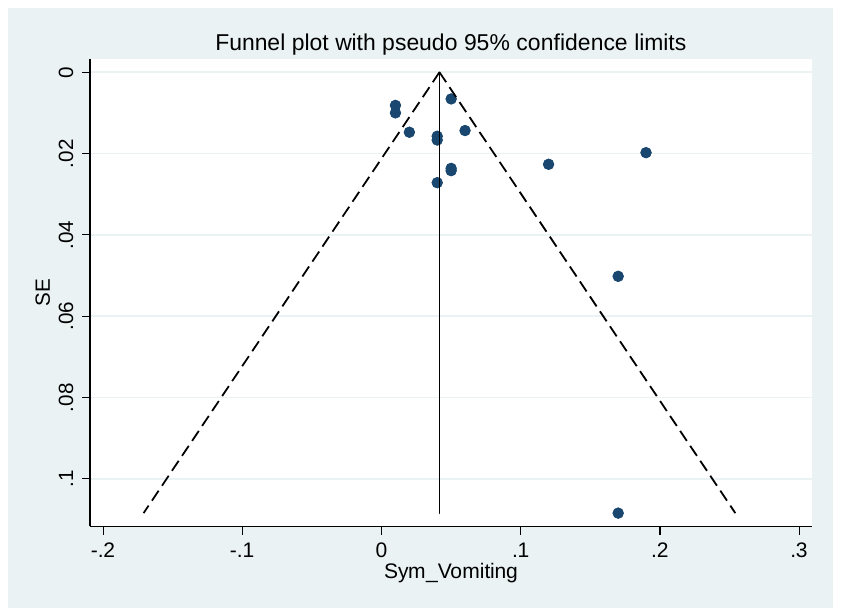

**S47. Abdominal pain**

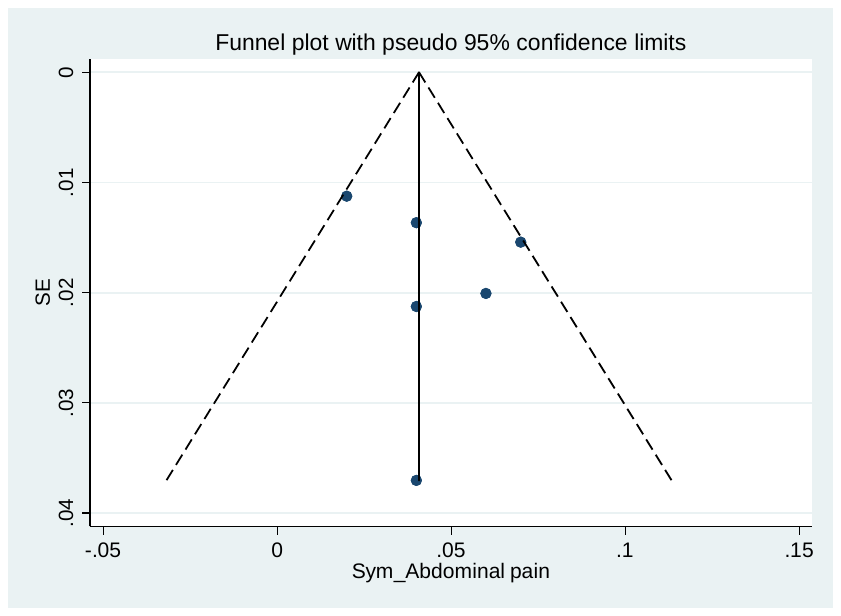

**S48. Dizziness**

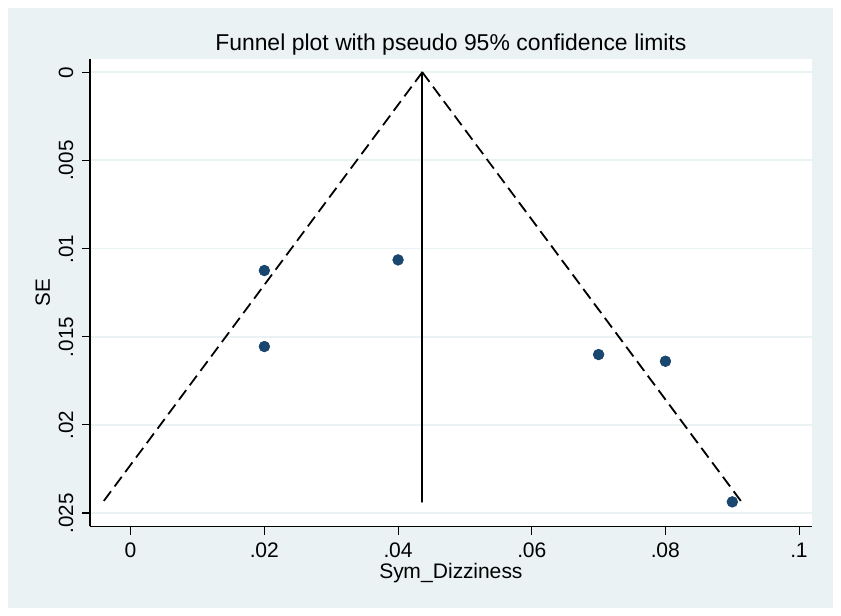

**S49. Anorexia**

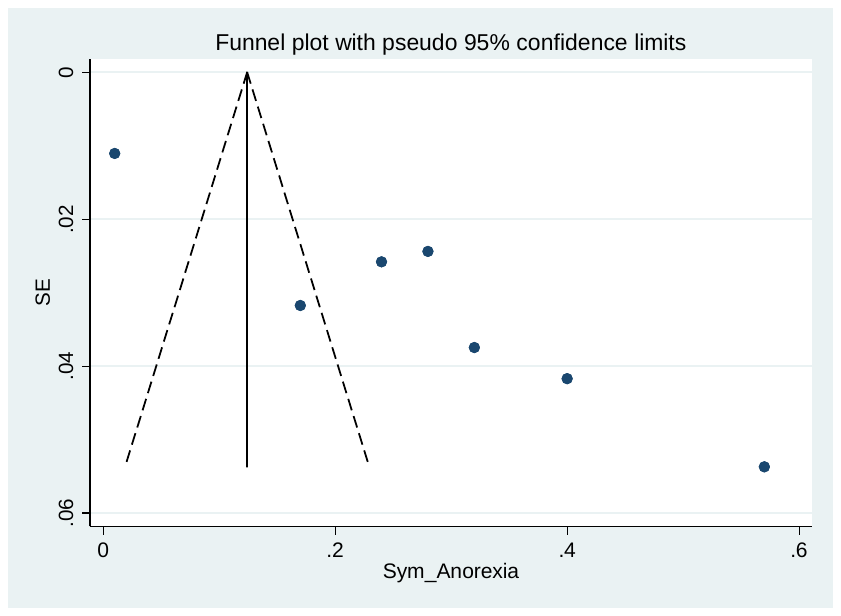

**S50. Pharyngalgia**

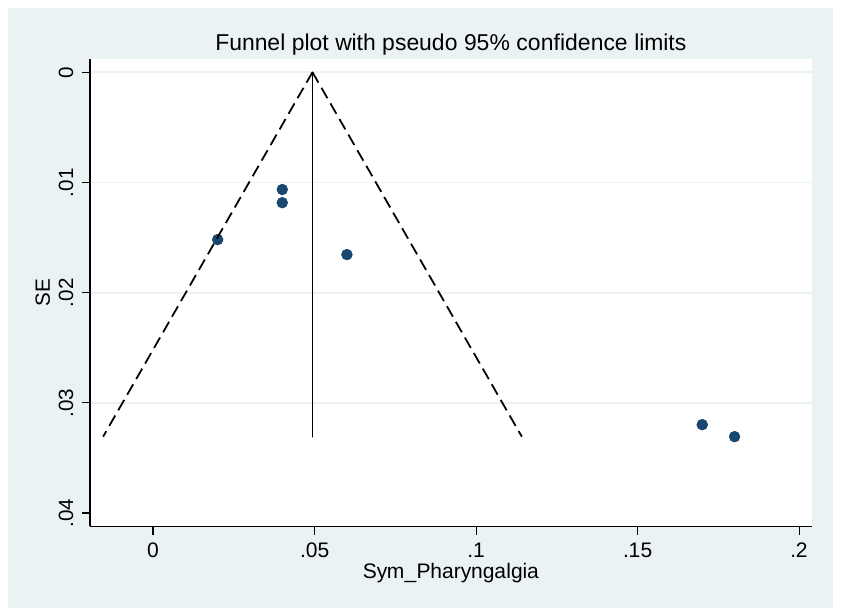

**S51. Haemoptysis**

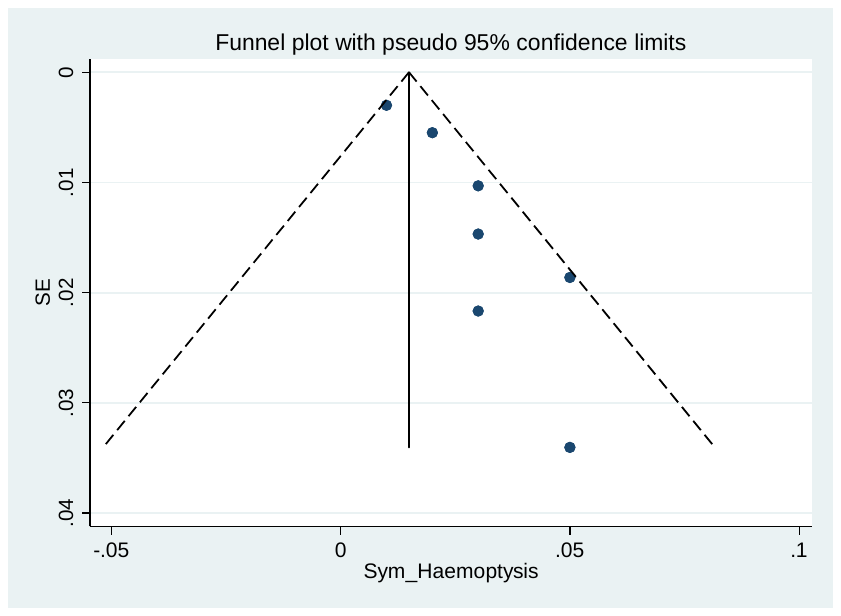

**Comorbidity**

**S52. Diabetes**

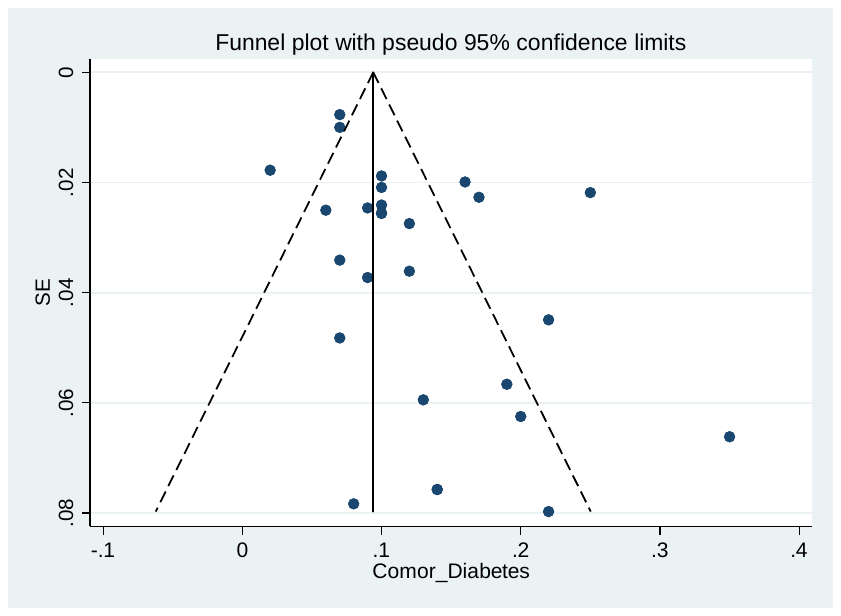

**S53. Hypertension**

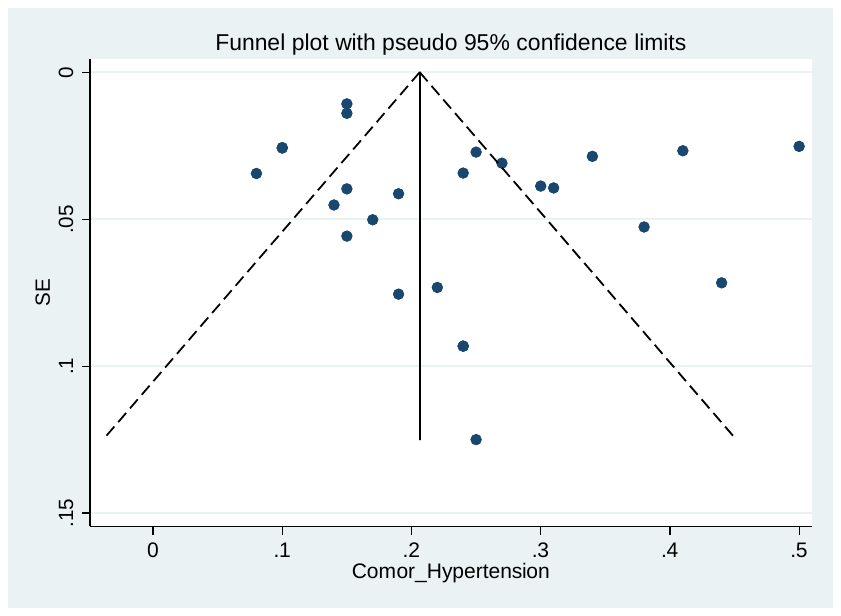

**S54. Cardiovascular disease**

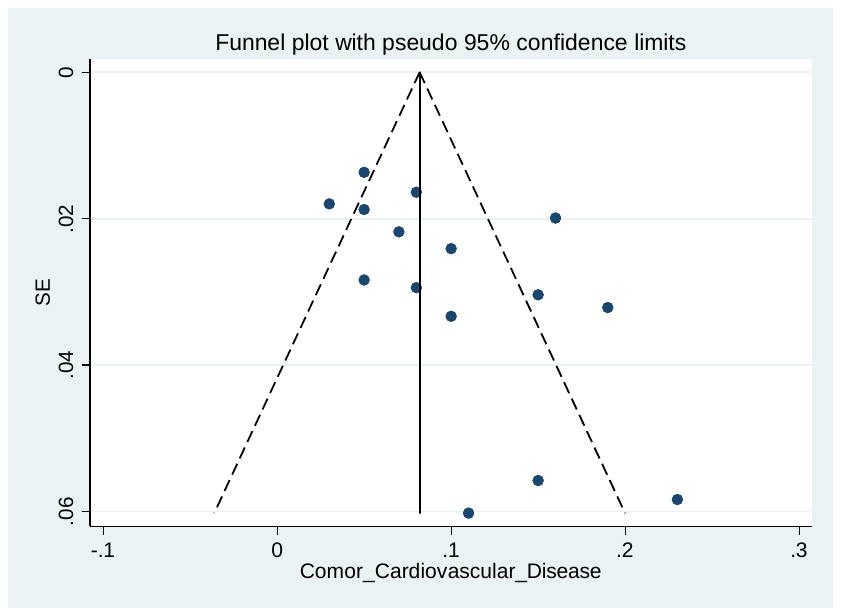

**S55. Coronary heart disease**

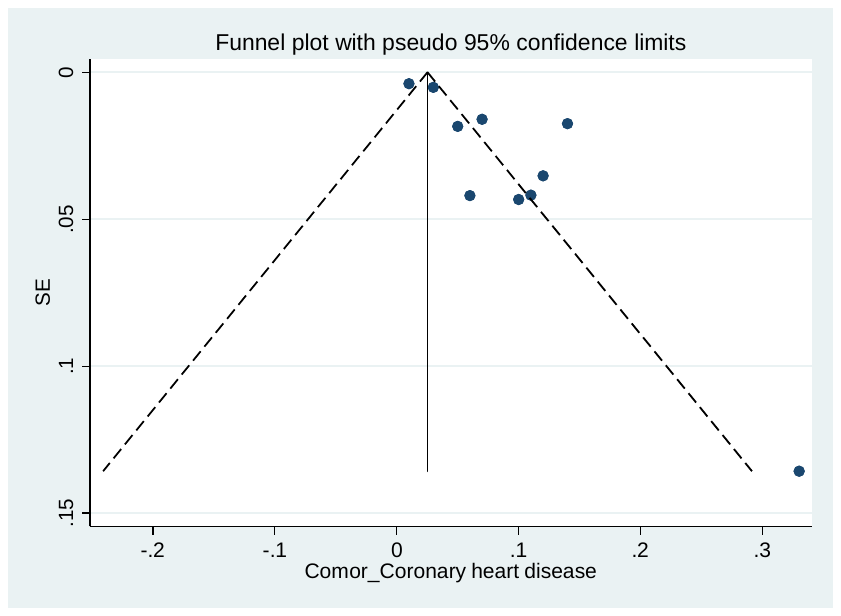

**S56. Cerebrovascular disease**

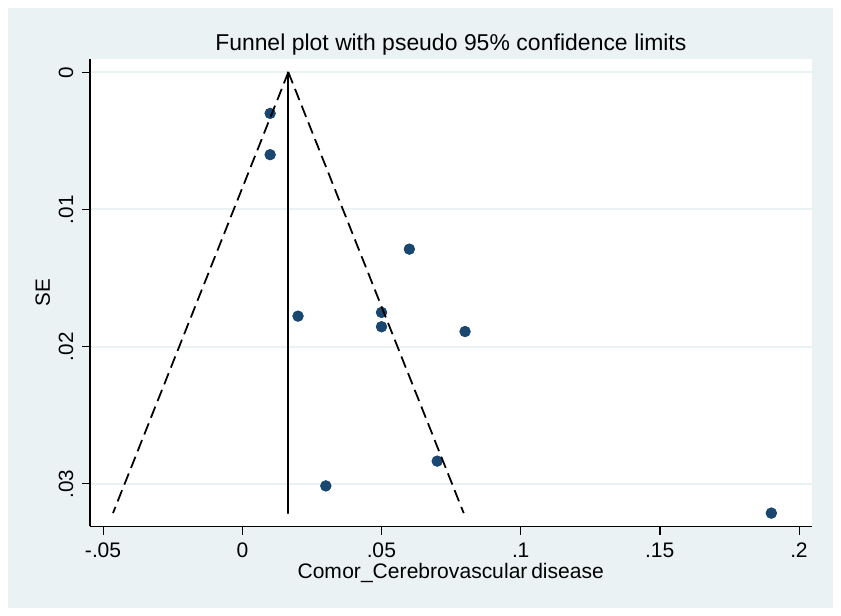

**S97. COPD / Lung disease**

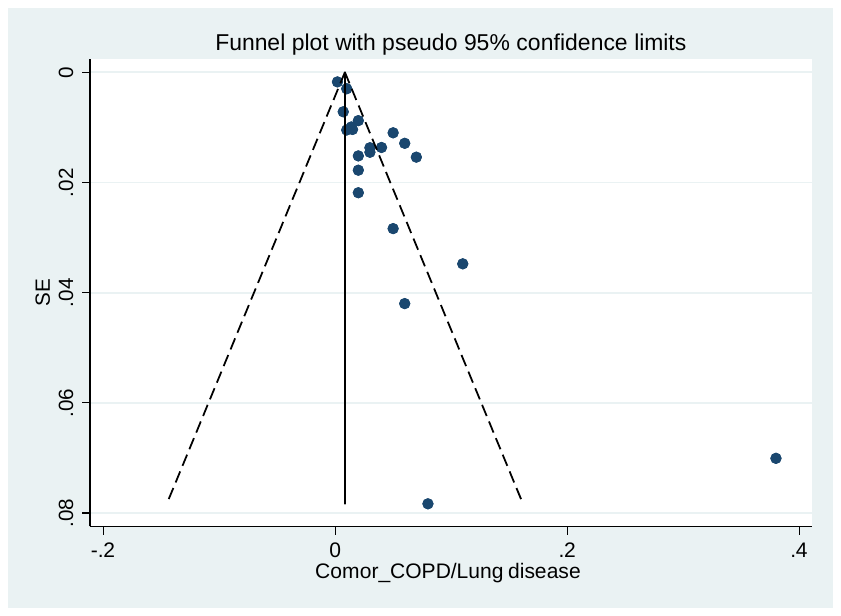

**S58. Chronic Liver disease**

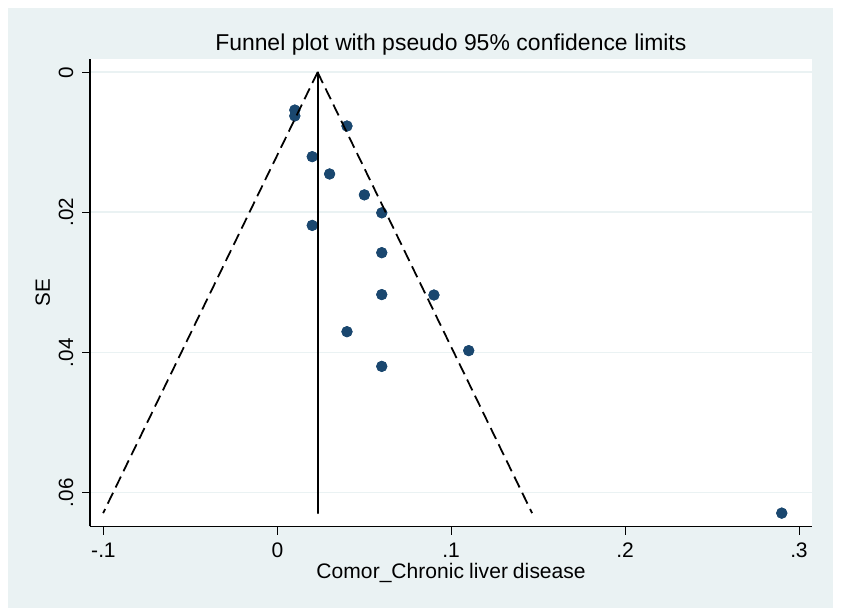

**S59. Chronic Renal disease**

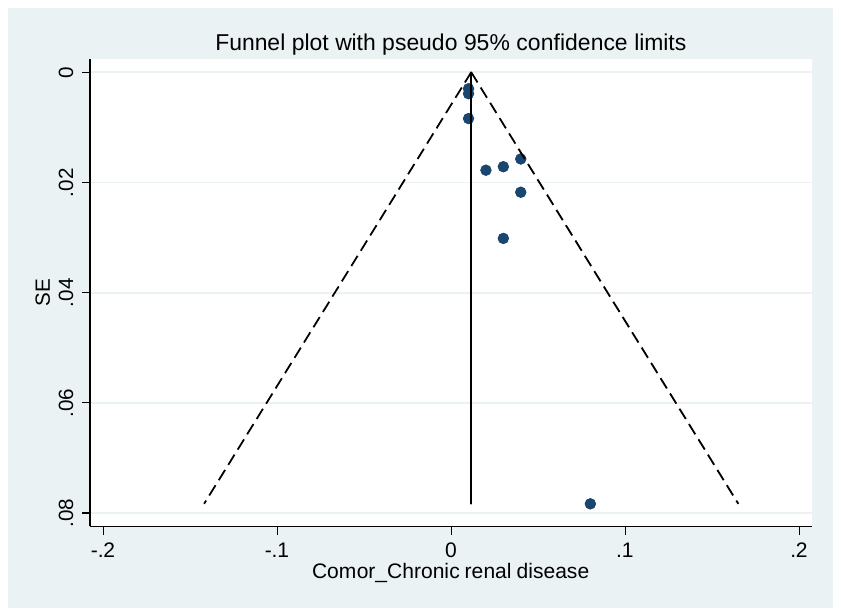

**S60. Chronic Kidney disease**

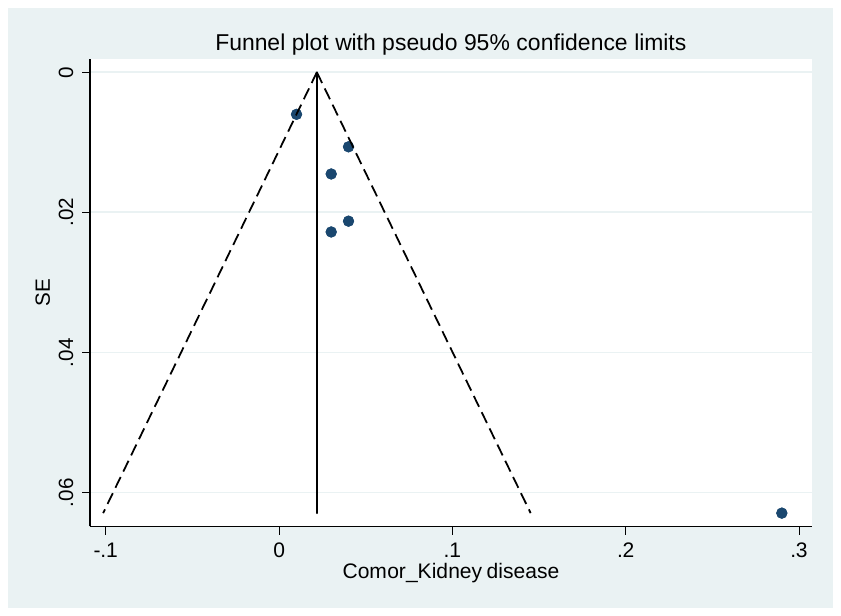

**S61. Malignancy**

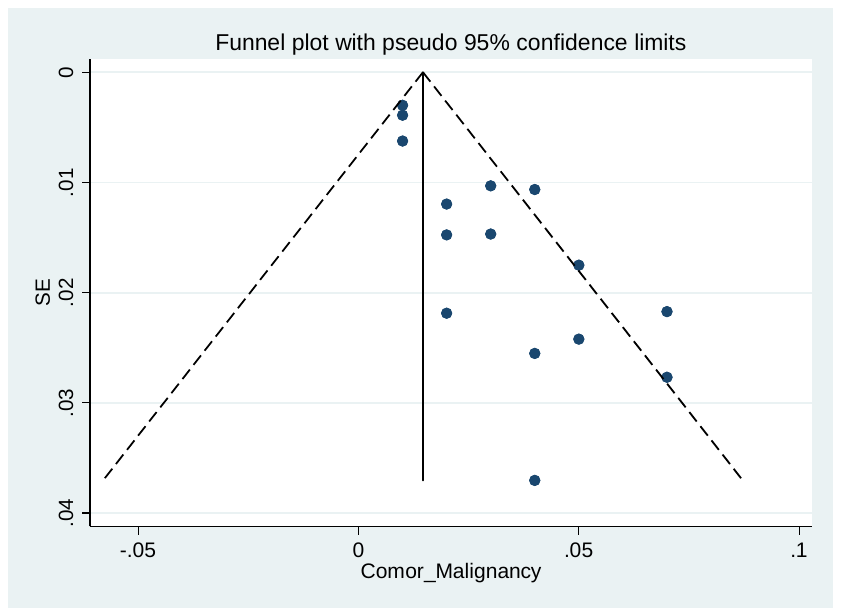

**S62. ARDS**

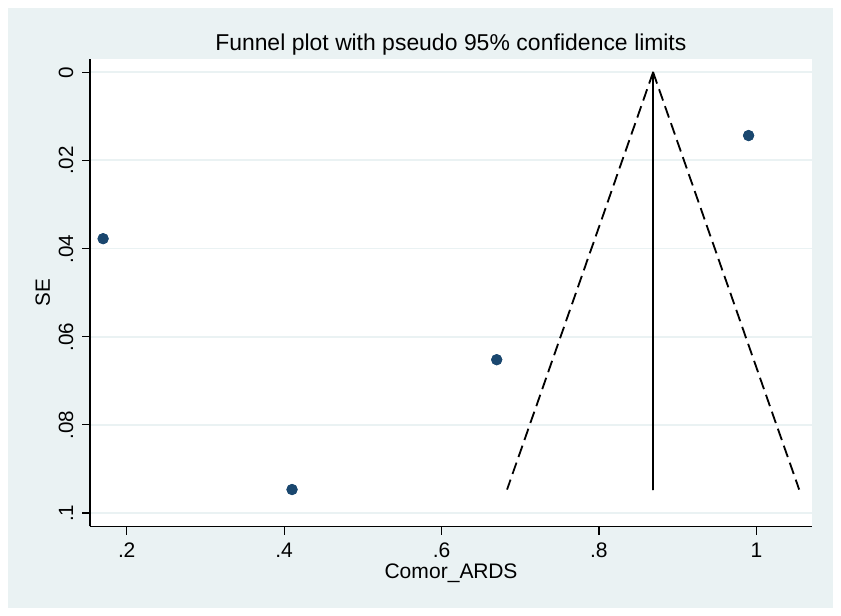
